# COVID-19: Time-Dependent Effective Reproduction Number and Sub-notification Effect Estimation Modeling

**DOI:** 10.1101/2020.07.28.20164087

**Authors:** Eduardo Atem de Carvalho, Rogerio Atem de Carvalho

**Affiliations:** Advanced Materials Laboratory, Universidade Estadual do Norte Fluminense, Brazil; Innovation Hub, Instituto Federal Fluminense, Brazil

**Keywords:** COVID-19, Pandemics, Infection Control, Models, Theoretical, Longitudinal Studies, Statistical Modeling, Epidemic Cycles

## Abstract

**Background:** Since the beginning of the COVID-19 pandemic, researchers and health authorities have sought to identify the different parameters that govern their infection and death cycles, in order to be able to make better decisions. In particular, a series of reproduction number estimation models have been presented, with different practical results.

**Objective:** This article aims to present an effective and efficient model for estimating the Reproduction Number and to discuss the impacts of sub-notification on these calculations.

**Methods:** The concept of Moving Average Method with Initial value (MAMI) is used, as well as a model for R_t_, the Reproduction Number, is derived from experimental data. The models are applied to real data and their performance is presented.

**Results:** Analyses on R_t_ and sub-notification effects for Germany, Italy, Sweden, United Kingdom, South Korea, and the State of New York are presented to show the performance of the methods here introduced.

**Conclusions:** We show that, with relatively simple mathematical tools, it is possible to obtain reliable values for time-dependent, incubation period-independent Reproduction Numbers (R_t_). We also demonstrate that the impact of sub-notification is relatively low, after the initial phase of the epidemic cycle has passed.

## 1) Introduction

The attempt to describe the different epidemic cycles that represent the current COVID-19 pandemic often comes up against the quality of the publicized data. For a number of practical reasons registration of deaths and of infections are inevitably imprecise, although can be corrected over time, as we discuss in [1]. This previous work suggests that one way around this effect is to apply the so-called Moving Average Method (MAM), in which the daily value of deaths is replaced by the sum of the current daily value with the values for the following six days, divided by seven. In other words, in a 7-day week, starting on Sunday and ending on Saturday, the average of the seven values will be computed in the Sunday value. This is the case with the Moving Average Method with Initial Value (MAMI). In the series analysis, the MAMI is seen as a moving window in time, starting from the first event (infection or death) to the last one. While in [1] we dealt with finding patterns in the number of deaths cycles, in order to predict the end of these cycles, in this paper we introduce a method to estimate the transmission rate or, in technical terms, the Basic Reproduction Number (R_0_), which in fact, we consider that varies in time, therefore it should be considered as a Transmission Function. Many works focused and continue to focus on establishing R_0_, the most recent listed on PubMed database (https://pubmed.ncbi.nlm.nih.gov/), with direct relationship with our work, are briefly commented as follows.

Musa et al. [2] estimate the exponential growth rate and basic reproduction number (R_0_) of COVID-19 in Africa to show the potential of the virus to spread. The authors analyzed the initial phase of the epidemic in Africa by using the simple exponential growth model. The Poisson likelihood framework is adopted for data fitting and parameter estimation, and the distribution of COVID-19 generation interval is modeled as Gamma distributions to compute the basic reproduction number. With a calculated R_0_, the authors quantified the instantaneous transmissibility of the outbreak by the time-varying effective reproductive number to show the potential of COVID-19 to spread across African region.

Hao et al. [3] reconstruct the full-spectrum dynamics of the COVID-19 outbreak between January 1 and March 8, 2020 in Wuhan, China, across five time periods based on key events and interventions. Following specific assumptions, the authors assumed that the transmission rate and the ascertainment rate did not change in the first two of the periods, and estimated rates across periods by Markov Chain Monte Carlo (MCMC) and further converted the transmission rate into the effective reproduction number R_e_. Using confirmed cases exported from Wuhan to Singapore, the authors conservatively estimated the ascertainment rate during the early outbreak in Wuhan. Sensitivity analyses to test the robustness was applied to an specific outlier data point, varying lengths of latent and infectious periods, duration from symptom onset to isolation, ratio of transmissibility of unascertained cases to ascertained cases, and the initial ascertainment rate.

Ogden et al. [4] introduce a two-folded approach with an agent-based model and a deterministic compartmental model. Both are Susceptible-Exposed-Infectious-Recovered (SEIR) type models with elements to model COVID-19 and impacts of Non-pharmaceutical interventions (NPIs). Parameters in the models are calibrated daily by literature searches, to ensure that evolving knowledge is captured by the models. The main objective of the modeling approaches was to compare the impacts of different NPIs, by adjusting Gaussian-shaped curves according to the parameters obtained daily and checking the R_e_.

Fawad et al. [5] aim at obtaining precise estimates of COVID-19 Pandemic current and future trends, by retrieving data from the Health Commission of Hubei, China, using Logistic-S curve model to estimate the current and future trends among 16 cities of Hubei, China from the 11^th^ of January to the 24^th^ of February, 2020. Mean Absolute Percentage Error (MAPE), Mean Absolute Deviation (MAD), and Mean Squared Deviation (MSD) were used to evaluate the predicted models for each city.

Lin et al. [6] aim to summarize the mathematical models that have been developed to understand and predict the infectiousness of COVID-19 to inform efforts to manage the outbreak in China. The authors searched various article bases for relevant studies published between 1^st^ December, 2019 and 21^st^, February, 2020, as well as built a Gaussian simulation for the evolution of the epidemic in Wuhan specifically. When R_0_ and R_c_ were stratified by region, differences among the two parameters remained statistically significant across all four studied Chinese regions.

Although the works listed above are quite recent, they are based on data from the beginning of the pandemic, and as we noted in [1], these may have led to later models that failed to make predictions. In fact, other authors identify deficiencies in the prevalent pandemic prediction models. Hon and Li [7] work on top of the Susceptible-Infectious-Removed (SIR) model and its variants for modeling the pandemic. The author however list that time-independent parameters in the classical models may not capture the dynamic transmission and removal processes, governed by virus containment strategies taken at various phases of the epidemic; and few models account for possible inaccuracies of the reported cases. The authors in [7] propose a Poisson model with time-dependent transmission and removal rates to account for possible random errors in reporting and estimate a time-dependent disease reproduction number, which may reflect the effectiveness of virus control strategies.

Similar problems with the early models on COVID-19 cycles were also reported by [8]. The authors present an analysis of these early models, mostly based on the city of Wuhan original outbreak. They present a statistical framework for comparing and combining different estimates of R_0_ (the transmission rate or reproductive number) across a wide range of models by decomposing the basic reproductive number into three key quantities: the exponential growth rate, the mean generation interval and the generation-interval dispersion. They apply their framework to early estimates of R_0_ for the SARS-CoV-2 outbreak, showing that many R_0_ estimates were overly confident. The results emphasize the importance of propagating uncertainties in all components of R_0_, including the shape of the generation-interval distribution, in efforts to estimate R_0_ at the outset of an epidemic. In general, the authors in [8] conclude that the models derived from the initial Chinese data resulted in low quality later predictions, in the same way that we point out in [1].

Therefore, there is room for proposing new forms of estimating R_0_ more precisely. Thus, this article, while recognizing the important contribution made by the other authors who preceded it, seeks simultaneously to propose a model that corrects their deficiencies and at the same time is simple to apply. Simplicity, combined with efficiency, is essential for its use to be disseminated by the health authorities of micro-regions and cities, since in [1] we propose that it is through the analysis of regional data that a better understanding of the patterns of the epidemic cycles is achieved.

## 2) Data Conditioning and Epidemic Cycle Definition

In the same way that was done in [1], in this work certain definitions have to be established before going further. Initially, we submit daily infection cases figures to the previously discussed Moving Average Method with Initial value (MAMI), and the critical epidemic cycle duration is defined by the time spent between its beginning, the peak and its closure. Also, it is considered that the cycle does start when the number of daily infections start to climb continuously past a threshold of 5% of the MAMI peak. The same criteria is used to define the end of the cycle, however, in this case when figures descent past 5% of the peak value. A False Peak is a data value that does not fit in the general cycle tendency. A Peak, or a True Peak, is the point where the cycle reverses itself from climbing to descending [1]. Finally, there is the question of terminology: we consider that there is an effective transmission rate that is dependent of time, represented by a function R_e_(t), however, for the sake of simplicity, it will be referenced using the usual terminology and representation of the area, which is simply R_t_.

### 2.1) MAMI Effect Over Reproduction Numbers

The impact of MAMI applied to registered numbers can be better understood by analyzing Figure 1. It is easily seems that MAMI bears greatest effect at the very beginning of the epidemic cycle, after a brief time, average and actual data tend to yield to the same value as cycles progress. It will be shown along this paper that the Reproduction Number varies most at the early stages and the use of MAMI is plainly justified to avoid numbers that are registered in batches and not into a smooth, daily, fashion.

**Figure 1.**
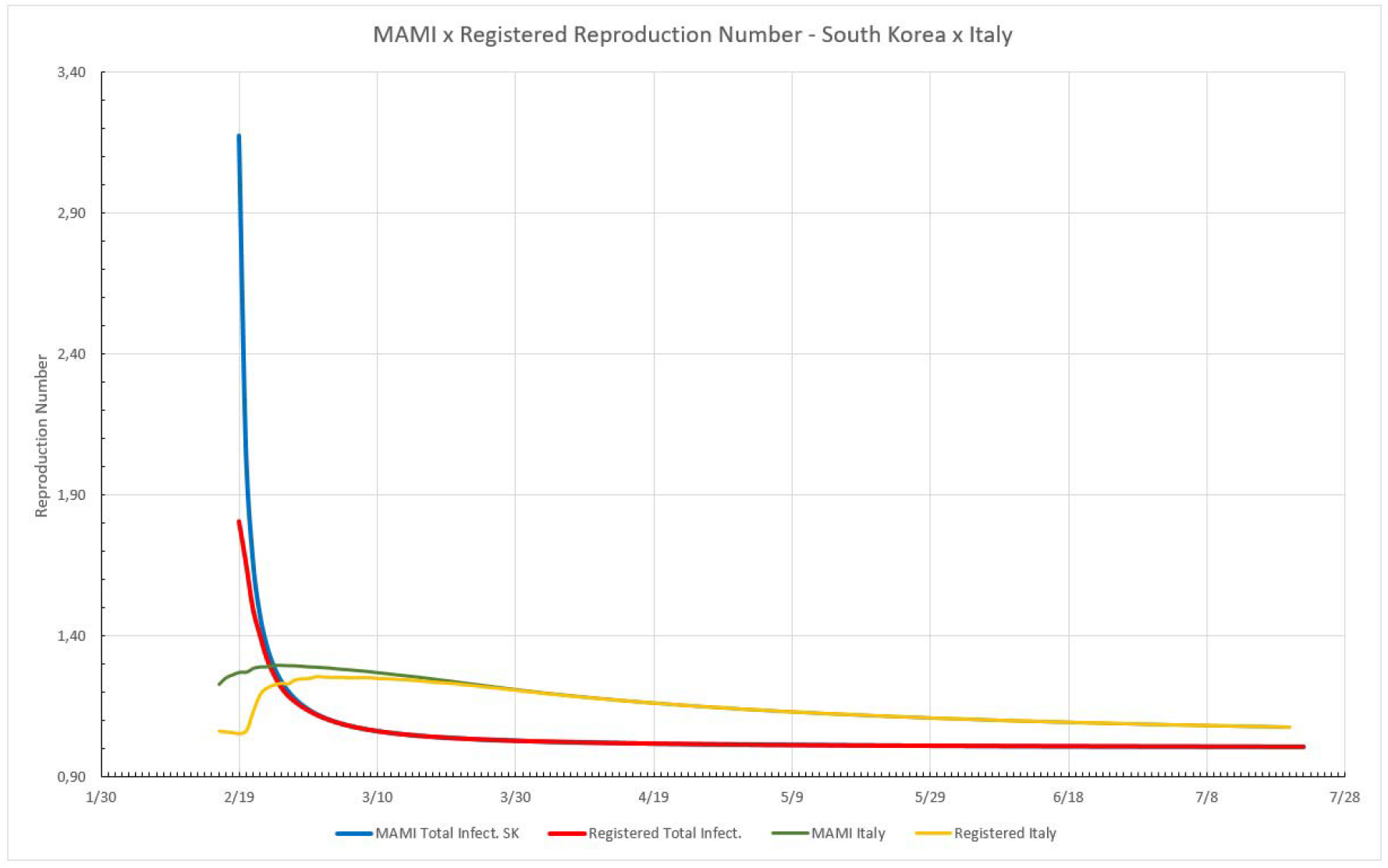
MAMI effect over Reproduction Numbers expressed for two different countries, South Korea and Italy. South Korea: the blue line is R_t_ obtained from MAMI applied to registered data, the red line is R_t_ determined for registered data. For Italy: the yellow line is R_t_ for registered data and the green line for MAMI applied to registered data.

### 2.2) The Progressive Sum of Infection Cases

The use of Moving Average Method, fixing the average of the seven days cycle value at the first day, as explained in [1], is used along this work. Daily values for total cases were collected from John Hopkins University’s website (https://coronavirus.jhu.edu/map.html) on the 22^sd^ of July, 2020. The number of infected should not be used straight as reported daily by local authorities, for reasons previously explained [1], but before, submitted to MAMI, and then applied to the described method. This method may be used for Progressive Sum of Infection Cases (PSIC), where the next day is always greater than the previous (or at least equal). In this case it is assumed that the whole number of infected is capable of contaminating and not only those listed on the day before, therefore is more realistically to use the summation of cases than the daily cases solely.

### 2.3) Reported Data

Germany is a Western country that was reported as exemplary in terms of application of NPIs, therefore it is going to be used as a starting point for this work. In Figure 2, the black line represents the number of daily cases, as listed at JHU’s site. Blue bars are these numbers submitted to MAMI and the red line represents the total cases to date and uses the right hand axis as reference.

**Figure 2.**
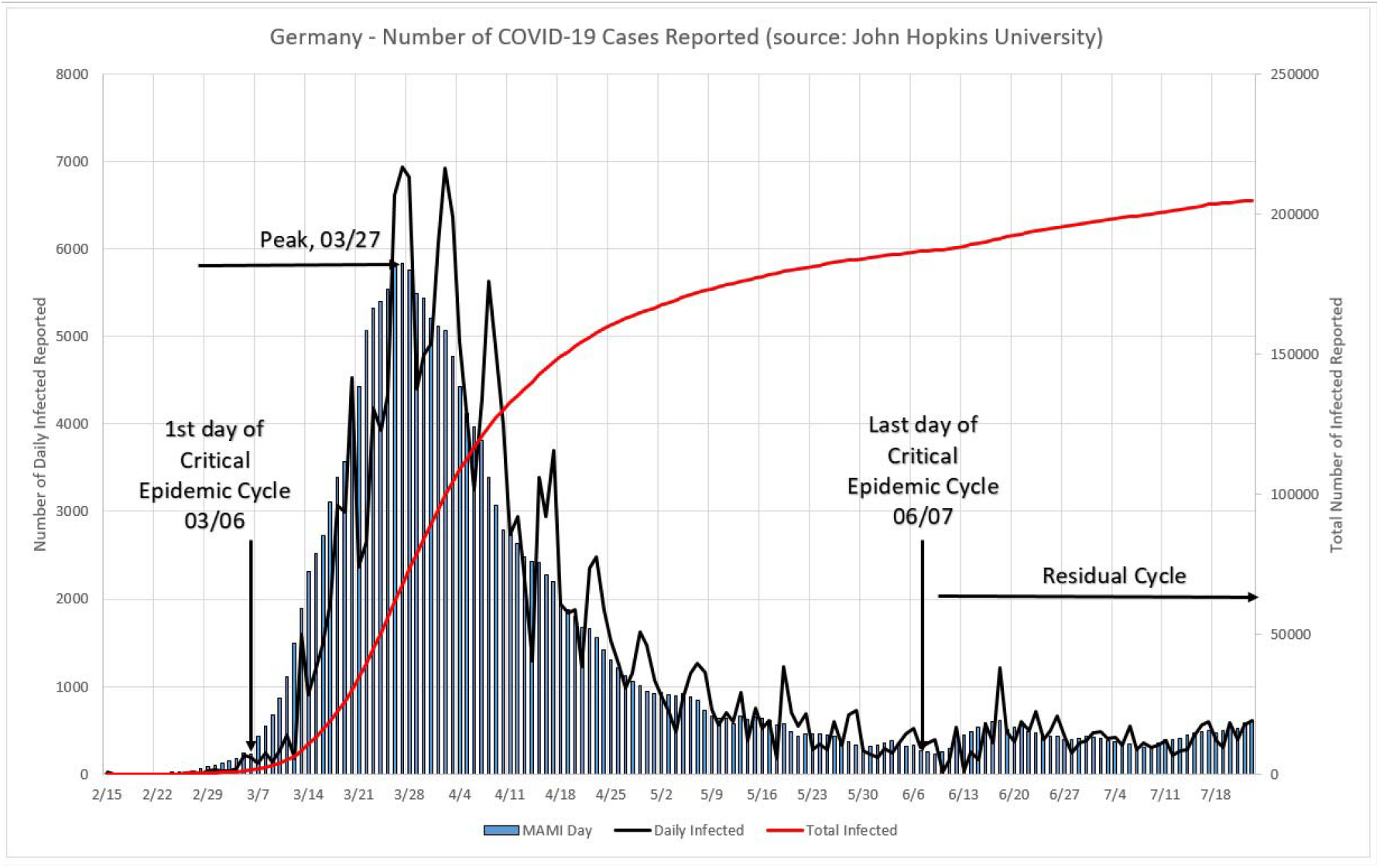
Number of COVID-19 cases reported for Germany. The black line represents the daily reported numbers, blue bars their MAMI, and the red line the total cases to date and uses the right hand axis as reference.

The same data source, calculation method, and display procedure is used for representing data from Italy, Sweden, UK, South Korea, and the State of New York, as presented in Figures 3 to 7. All data were collected on the 22^nd^ of July, 2020.

**Figure 3.**
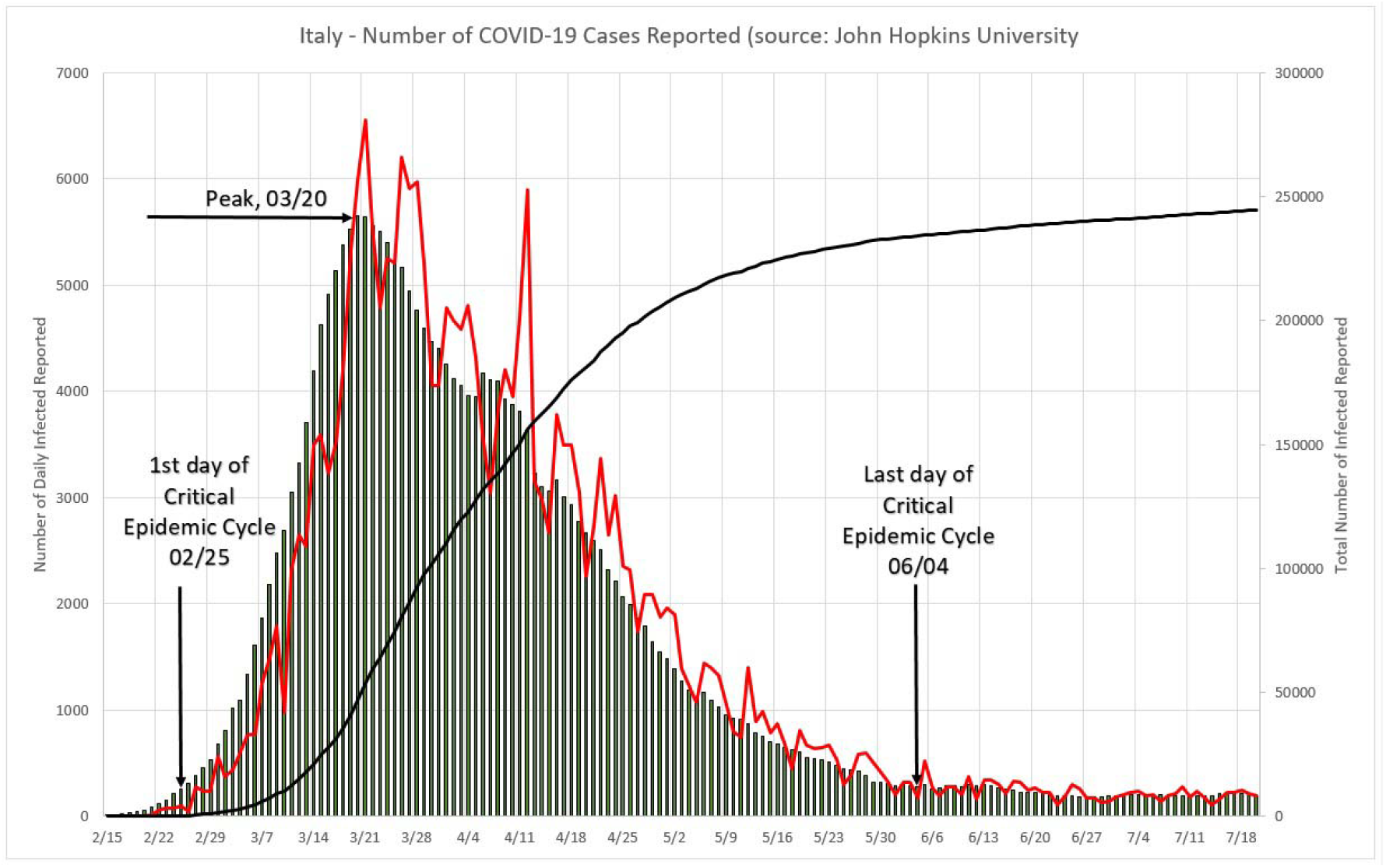
Number of COVID-19 cases reported for Italy. The black line represents the daily reported numbers, blue bars their MAMI, and red line the total cases to date and uses the right hand axis as reference.

Table 1 displays the basic critical cycles numbers presented in this work. Note that Sweden and New York State still have not closed their critical cycles, if a 5% percent limit criteria is used. It does contain the dates for beginning and end of the critical cycle, its duration, and the peak day. The maximum value of daily cases after MAMI was applied to registered daily cases and the 5% limit used to determine the cycle boundaries.

**Table 1.** Basic critical cycles numbers

| Place | Cycle Starts | Cycle Ends | Duration | Peak | MAMI Max | 5% Limit |
| --- | --- | --- | --- | --- | --- | --- |
| Germany | 03/06 | 06/07 | 93 | 03/27 | 5837 | 292 |
| Italy | 02/25 | 06/04 | 100 | 03/20 | 5651 | 283 |
| Sweden | 03/04 | -- | -- | 06/23 | 1248 | 62 |
| UK | 03/06 | 06/27 | 93 | 04/16 | 4868 | 243 |
| South Korea | 02/15 | 04/09 | 54 | 02/26 | 623 | 31 |
| NY State | 03/13 | -- | -- | 04/04 | 10059 | 593 |

**Table 2.** lists the errors associated with ignoring the existence of Sub Notification into the epidemic cycle.

| Sub Notif. | Max Error (%) | Min. Error (%) | Days Until ≤ 5% | Error (%) at Peak Day |
| --- | --- | --- | --- | --- |
| 3x | 5.34 | 0.97 | 2 | 2.64 |
| 5x | 7.73 | 1.41 | 12 | 3.85 |
| 10x | 10.87 | 2.02 | 25 | 5.46 |
| 15x | 12.66 | 2.37 | 33 | 6.39 |
| 20x | 13.91 | 2.62 | 39 | 7.05 |
| **25x** | 14.87 | 2.81 | 43 | 7.55 |
| **30x** | 15.64 | 2.97 | 47 | 7.96 |
| **40x** | 16.84 | 3.21 | 52 | 8.60 |

**Table 3.** List the estimated errors associated with ignoring the existence of Sub Notification into the epidemic cycle in South Korea.

| Sub Notif. | Max Error (%)<br>– First Day | Min. Error (%)<br>– Last Day | Days Until ≤<br>5% | Error (%) at<br>Peak Day |
| --- | --- | --- | --- | --- |
| **3x** | 4.47 | 1.40 | -- | 3.09 |
| **5x** | 6.49 | 2.04 | 21 | 4.49 |
| **10x** | 9.15 | 2.91 | 27 | 6.37 |
| **15x** | 10.67 | 3.41 | 29 | 7.45 |
| **20x** | 11.73 | 3.77 | 35 | 8.20 |
| **25x** | 12.55 | 4.04 | 39 | 8.79 |
| **30x** | 13.21 | 4.27 | 43 | 9.26 |
| **40x** | 14.25 | 4.62 | 48 | 10.00 |

**Table 4.** lists the errors associated with ignoring the existence of Sub Notification into the epidemic cycle - Italy.

| **Sub Notif.** | **Max Error (%)** | **Min. Error (%)** | **Days Until ≤ 5%** | **Error (%) at Peak Day** |
| --- | --- | --- | --- | --- |
| **3x** | 3.85 | 0.85 | -- | 2.09 |
| **5x** | 5.59 | 1.25 | 4 | 3.05 |
| **10x** | 7.89 | 1.78 | 17 | 4.33 |
| **15x** | 9.22 | 2.09 | 25 | 5.07 |
| **20x** | 10.15 | 2.31 | 31 | 5.60 |
| **25x** | 10.86 | 2.48 | 35 | 6.00 |
| **30x** | 11.44 | 2.62 | 39 | 6.33 |
| **40x** | 12.34 | 2.84 | 44 | 6.85 |

**Table 5.** lists the errors associated with ignoring the existence of Sub Notification into the epidemic cycle in Sweden.

| Sub Notif. | Max Error (%) | Min. Error (%)* | Days Until ≤ 5% | Error (%) at Peak Day |
| --- | --- | --- | --- | --- |
| **3x** | 5.92 | 0.69 | 4 | 0.85 |
| **5x** | 8.55 | 1.01 | 14 | 1.24 |
| **10x** | 12.01 | 1.45 | 27 | 1.77 |
| **15x** | 13.97 | 1.70 | 35 | 2.08 |
| **20x** | 15.33 | 1.88 | 41 | 2.30 |
| **25x** | 16.37 | 2.02 | 45 | 2.46 |
| **30x** | 17.22 | 2.13 | 49 | 2.60 |
| **40x** | 18.53 | 2.31 | 54 | 2.82 |

**Table 6.** lists the errors associated with ignoring the existence of Sub Notification into the epidemic cycle in NY State.

| **Sub Notif.** | **Max Error (%)** | **Min. Error (%)\*** | **Days Until ≤ 5%** | **Error (%) at Peak Day** |
| --- | --- | --- | --- | --- |
| **3x** | 11.49 | 0.78 | 13 | 3.48 |
| **5x** | 16.37 | 1.13 | 23 | 5.06 |
| **10x** | 22.57 | 1.62 | 36 | 7.16 |
| **15x** | 25.98 | 1.90 | 44 | 8.36 |
| **20x** | 28.31 | 2.10 | 50 | 9.21 |
| **25x** | 30.07 | 2.26 | 54 | 9.86 |
| **30x** | 31.47 | 2.38 | 58 | 10.39 |
| **40x** | 33.63 | 2.58 | 63 | 11.22 |

**Table 7.** lists the errors associated with ignoring the existence of Sub Notification into the epidemic cycle – the UK.

| **Sub Notif.** | **Max Error (%)** | **Min. Error (%)** | **Days Until ≤ 5%** | **Error (%) at Peak Day** |
| --- | --- | --- | --- | --- |
| **3x** | 5.10 | 0.82 | 1 | 1.76 |
| **5x** | 7.38 | 1.19 | 11 | 2.56 |
| **10x** | 10.38 | 1.70 | 24 | 3.65 |
| **15x** | 12.10 | 2.00 | 32 | 4.27 |
| **20x** | 13.29 | 2.21 | 38 | 4.72 |
| **25x** | 14.21 | 2.37 | 42 | 5.06 |
| **30x** | 14.95 | 2.51 | 46 | 5.34 |
| **40x** | 16.11 | 2.72 | 51 | 5.78 |

## 3) Daily Number of Infected

This paper proposes an approach that considers an effective, time-varying Reproduction Number, calculating its value by means of experimental data. Therefore, we seek to develop a mathematical model that is efficient and at the same time is fast computed. To achieve this objective, the cycle data were studied in detail and the following experimental formulations were developed.

The total number of infected daily (I_d_), during a period of time t, can be described as a function of the daily increase rate factor (1 + b) multiplied by a scale factor, as described in Formula (1):

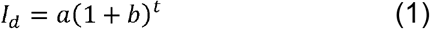

In Formula (1), “a” is the scale factor and “b” is the absolute daily increase rate, or instantaneous rate and is defined as:

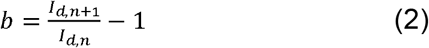

Where I_d,n+1_ is the current day and I_d,n_ the previous day.

Formula (1) can be written as:

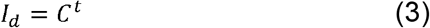

Where C is the the Time-Dependent Effective Reproduction Number, R_e_(t), or R_t_ for short, which is obtained from experimental data. For the Reproduction Number determination, it is necessary to determine the scale factor “a”. Therefore, “a” takes the following form:

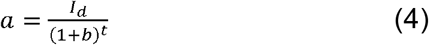

Finally, from Formulas (3) and (4):

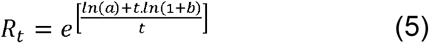

In order to map the interpretation proposed from Formulas (1) to (5) to the classical mathematical interpretation for the Reproduction Number, R_0_, an equivalence transformation will be described as follows.

From the classical definition of R_0_, let:

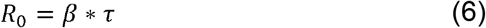

Where β is infection-producing contacts per unit time (instantaneous rate), with a mean infectious period of τ.

Formula (6) can be transformed into:

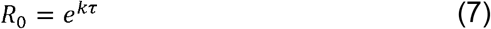

From Formulas (5) and (7) we have:

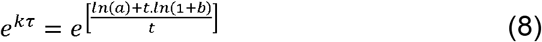

In (8) all dimensional units are compatible, therefore our transformations to obtain R_t_ in Formula (5) are valid. Formula (5) was obtained from experimental data, and it is at the core of the model here proposed. From this point onwards, R_t_ must be interpreted as R_e_(t) as explained before, in the interpretation of Formula (3).

During the analysis of data, we noted that the daily increase rate factor, (1 + b), is not enough to describe the number of contaminated cases registered at one given day, because it just informs the absolute increase ratio occurred from one day to the next. The Reproduction Number coefficient needs more numerical information in order to be able to express correctly the magnitude of daily numbers. It needs the scale factor “a” to bring more information on the phenomenon. As an example of this finding, Figure 7 shows that while the (1 + b) factor varies rapidly from one day to the R_t_, drops steadily, changing slowly as the exponential time grows. The same behavior is displayed by the total daily registered number of deaths, which keeps growing smoothly. This is the numerical evidence that the factor (1+b) alone cannot describe the total number of deaths.

**Figure 4.**
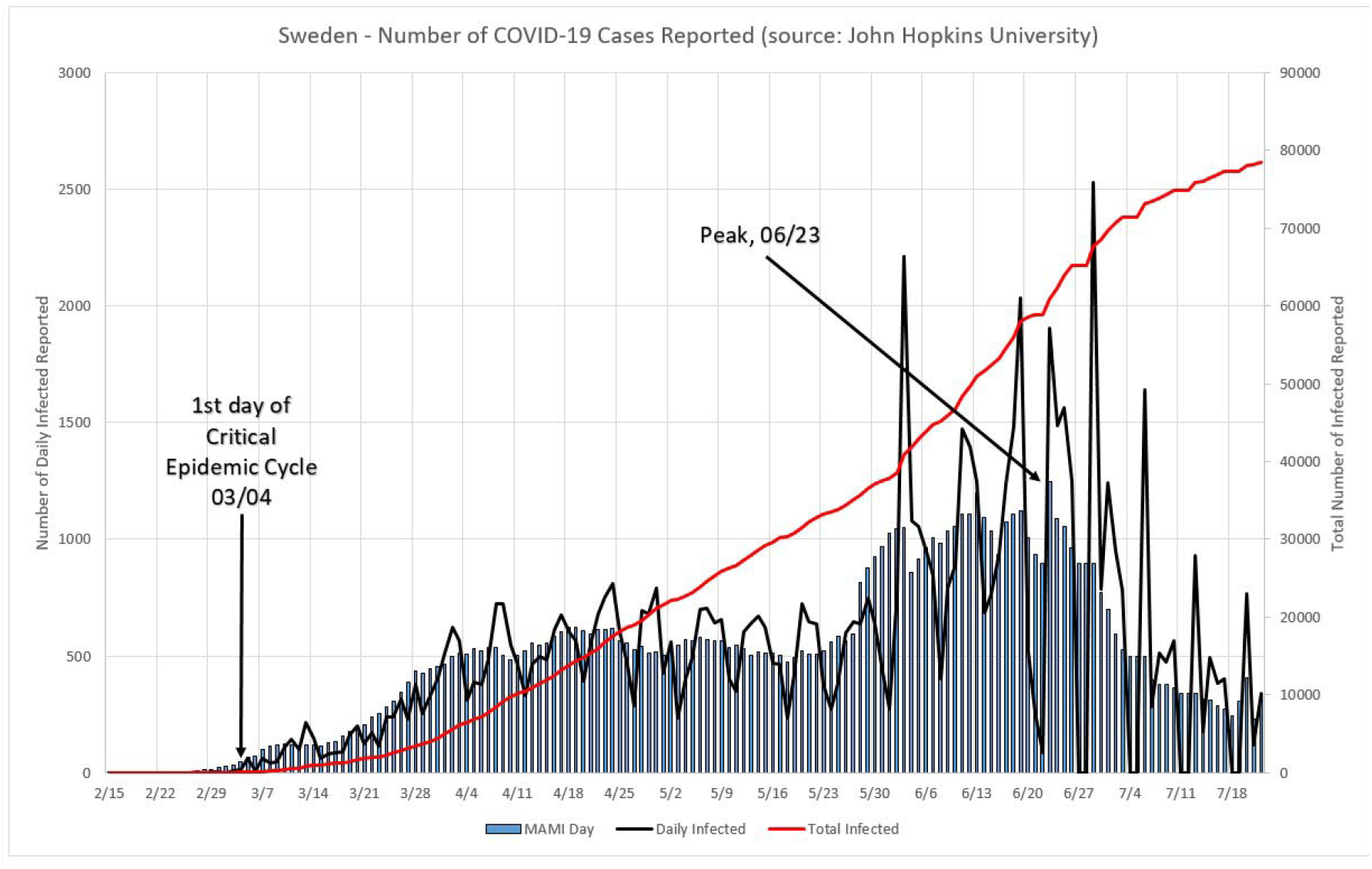
Number of COVID-19 cases reported for Sweden. The black line represents the daily reported numbers, blue bars their MAMI, and red line the total cases to date and uses the right hand axis as reference.

**Figure 5.**
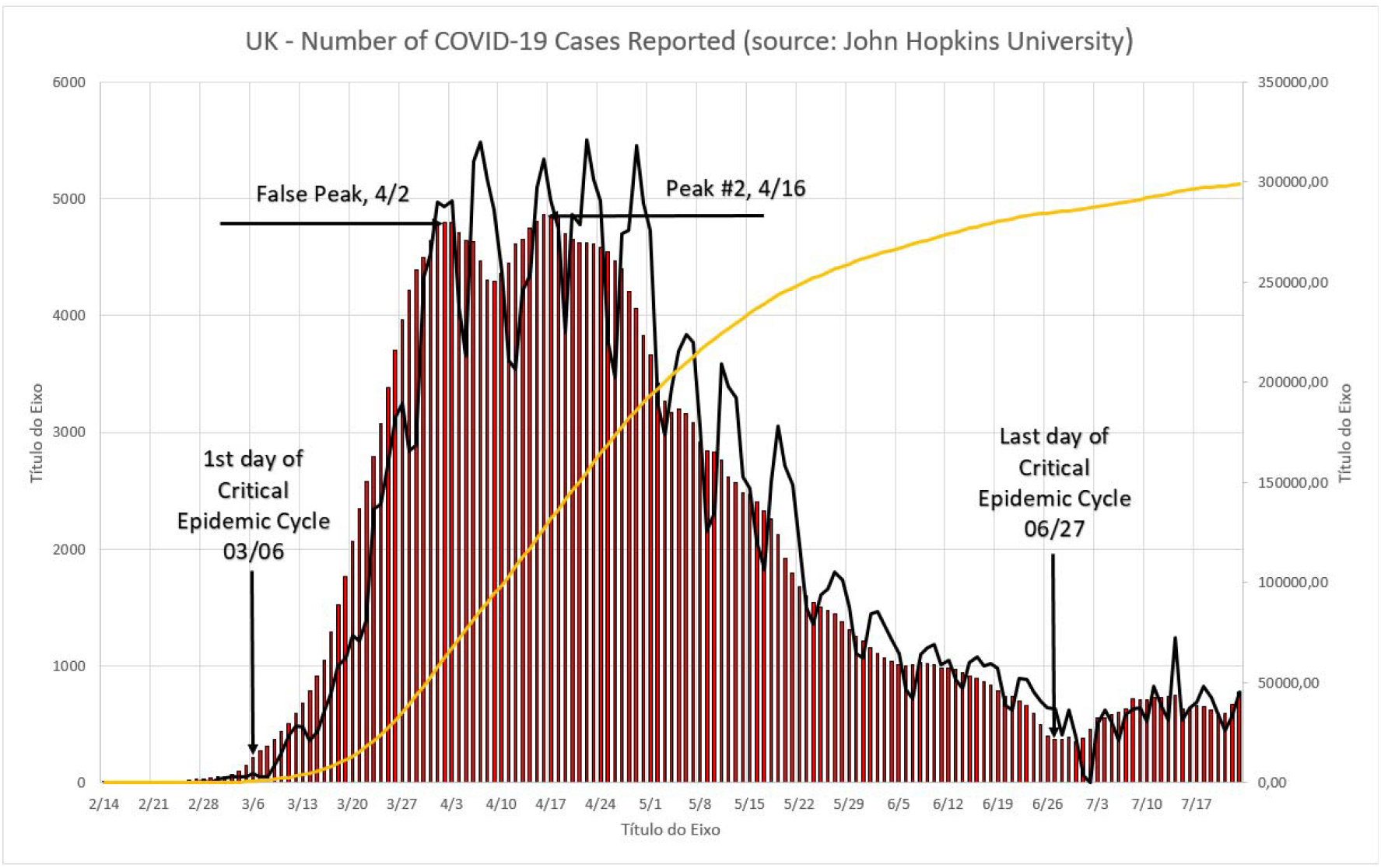
Number of COVID-19 cases reported for the UK. The black line represents the daily reported numbers, blue bars their MAMI, and red line the total cases to date and uses the right hand axis as reference.

**Figure 6.**
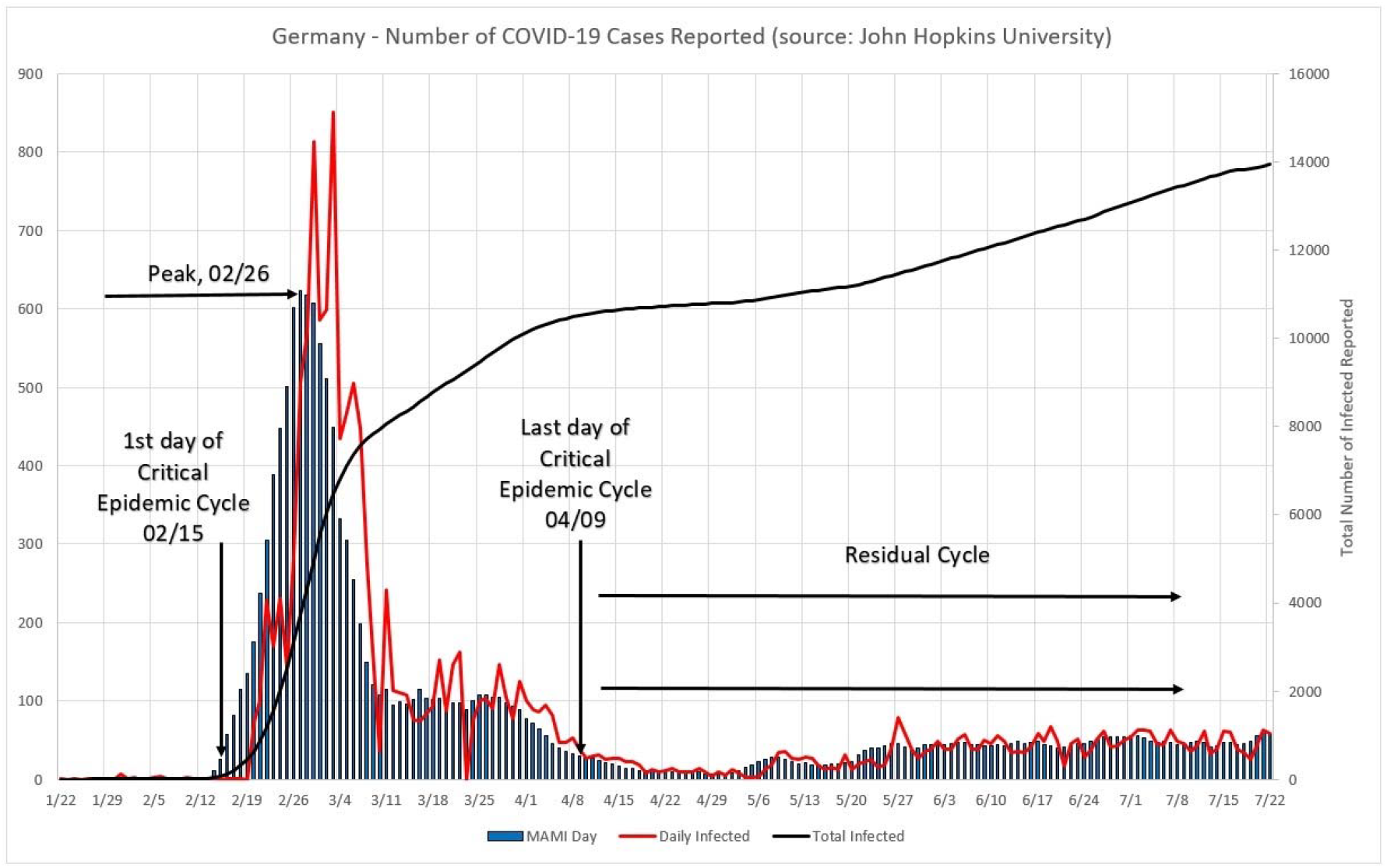
Number of COVID-19 cases reported for South Korea. The black line represents the daily reported numbers, blue bars their MAMI, and red line the total cases to date and uses the right hand axis as reference.

**Figure 7.**
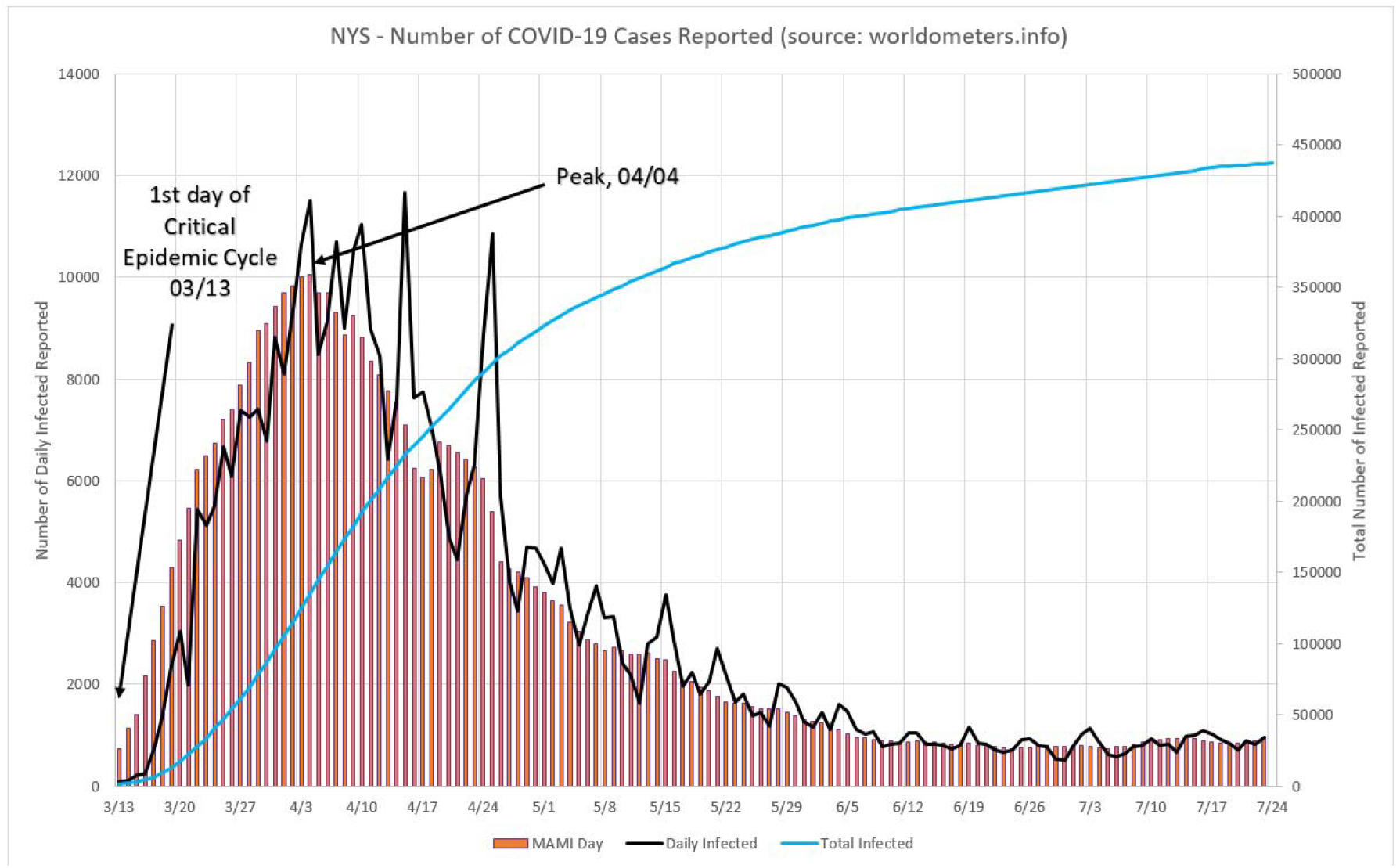
Number of COVID-19 cases reported for NY State. The black line represents the daily reported numbers, red bars their MAMI, and blue line the total cases to date and uses the right hand axis as reference.

**Figure 8.**
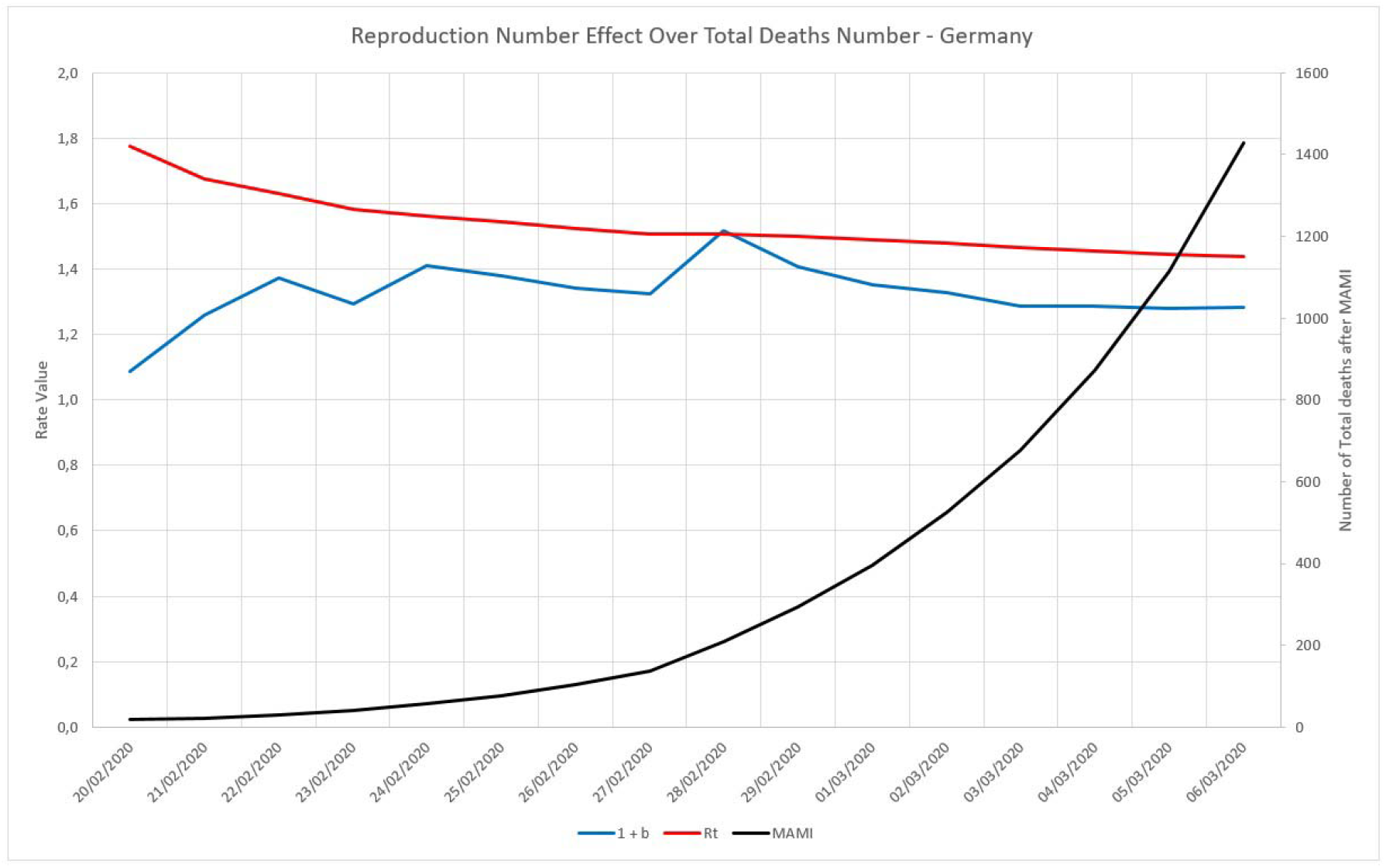
Behaviors of (1 + b) and R_t_ factors, for the first 20 days in the German epidemic cycle.

## 3.1) Impact of Incubation Period over R_t_

The expressions developed for this work in Formulas (1) to (5) do not explicitly take into account the so-called incubation period for the studied disease. The instantaneous rate of change, or daily increase in number of registered infected individuals is calculated as defined in Formula (5). For the sake of thoroughness, three simulations were ran, for a 5, 10 and 15 days incubation period. This was achieved by redefining expression (1+b) for a new set of parameters, basically dividing the total number of reported cases for a given day by the number registered in 5, 10 and 15 days before. This way, the term (1 + b) now would reflect the incubation period over R_t_. All simulations yielded zero (0%) change, to the fourth significant figure. Therefore, it is assumed that the described method is inherently insensitive to incubation period variations or influence, reinforcing its simplicity and robustness.

## 4) Reproduction Numbers: Case Studies

### 4.1) Germany

In Figure 9, three distinct zones are formed. Zone “a” is in the very beginning of the cycle and the Reproduction Number varies from 1.10 to 1.48 from one day to the next, this probably is just the reflection of large initial variation in numbers, but only if we limit this zone to no more than 5% of the MAMI peak value. It is easy to notice that the figures bear small influence in the overall disease behavior. Zone “b” describes transmissivity during the critical disease cycle (from March, the 6^th^ to June the 7^th^), where a rapid increase in daily cases stops only around the peak than drops steadily towards the end. This is the most lethal period of the epidemic cycle and it is considered over once a 5% peak level is reached again. The remaining time, Zone “c”, is the residual cycle that appears in all countries and places facing the COVID-19 crisis. In absolute values the Reproduction Number for the critical period starts with a 1.30 value and drops continuously towards 1.00, although never quite reaches it (as for the time of this paper was written).

**Figure 9.**
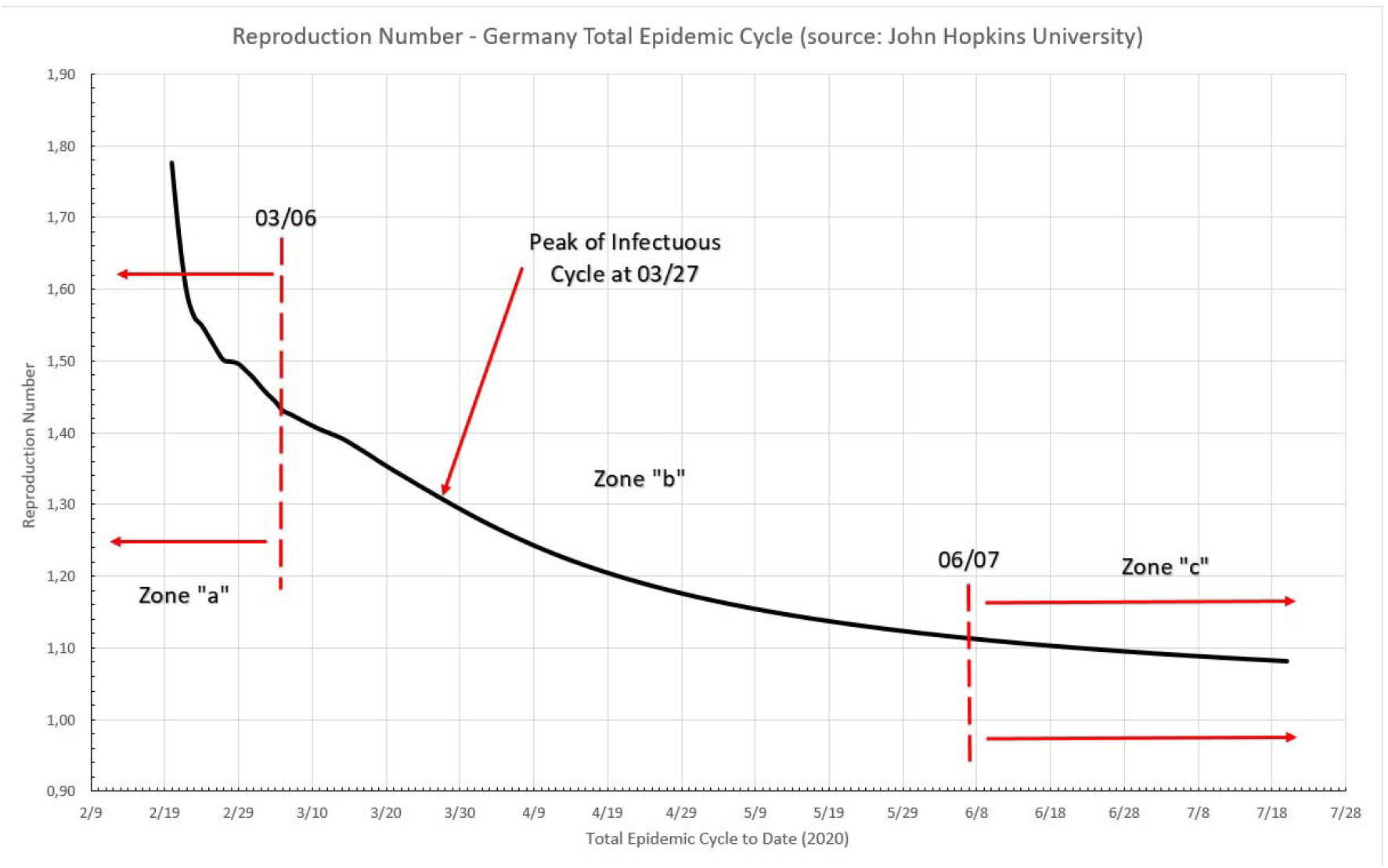
Total epidemic cycle in Germany, using the number of infected people daily.

### 4.2) Italy

It can be seem in Picture 10 that three distinct zones are formed. Zone “a” is in the beginning of the cycle and the Reproduction Number varies from 1.78 to 1.44 from one day to the next, once again this probably is just the reflection of large initial variation in number, but this zone is limited to no more than 5% of the MAMI peak value. It is easy to notice that the figures bear small influence in the overall disease behavior. Zone “b” describes transmissivity during the critical disease cycle (from February 25^th^ to June 15^th^). This is the most lethal period of the epidemic cycle and it is considered over once a 5% peak level is reached again. The remaining time, Zone “c”, is the residual cycle. In absolute values the Reproduction Number for the critical period starts with a 1.44 value and drops continuously towards 1.12.

### 4.3) Sweden

It can be seem in Picture 11 that two distinct zones are formed, once Sweden is considered, by the 5% criteria an “ongoing” epidemic cycle, although in the present date, close to the end. Zone “a” is in the beginning of the cycle and the Reproduction Number varies from circa 1.33 to 1.16 from one day to the next, once again this probably is just the reflection of large initial variation in number, but this zone is limited to no more than 5% of the MAMI peak value. It is easy to notice that the figures bear small influence in the overall disease behavior. Zone “b” describes transmissivity during the critical disease cycle (from March 4^th^ onwards). This is the most lethal period of the epidemic cycle and it is considered over once a below 5% peak level is reached again. In absolute values the Reproduction Number for the critical period starts with a 1.16 value and drops continuously towards 1.07.

### 4.4) United Kingdom

In Figure 12, two distinct zones are formed, because the UK critical cycle was considered over exactly at the data collection day. Zone “a” is in the beginning of the cycle and the Reproduction Number oscillates between 1.48 and 1.37 from one day to the next, once again this probably is just the reflection of large initial variation in number, but this zone is limited to no more than 5% of the MAMI peak value. Zone “b” describes transmissivity during the critical disease cycle (from March 6^th^ onwards, up to July, the 21^st^). This is the most lethal period of the epidemic cycle and it is considered over once a below 5% peak level is reached again. In absolute values the Reproduction Number for the critical period starts with a 1.37 value and drops continuously towards 1.08.

**Picture 10.**
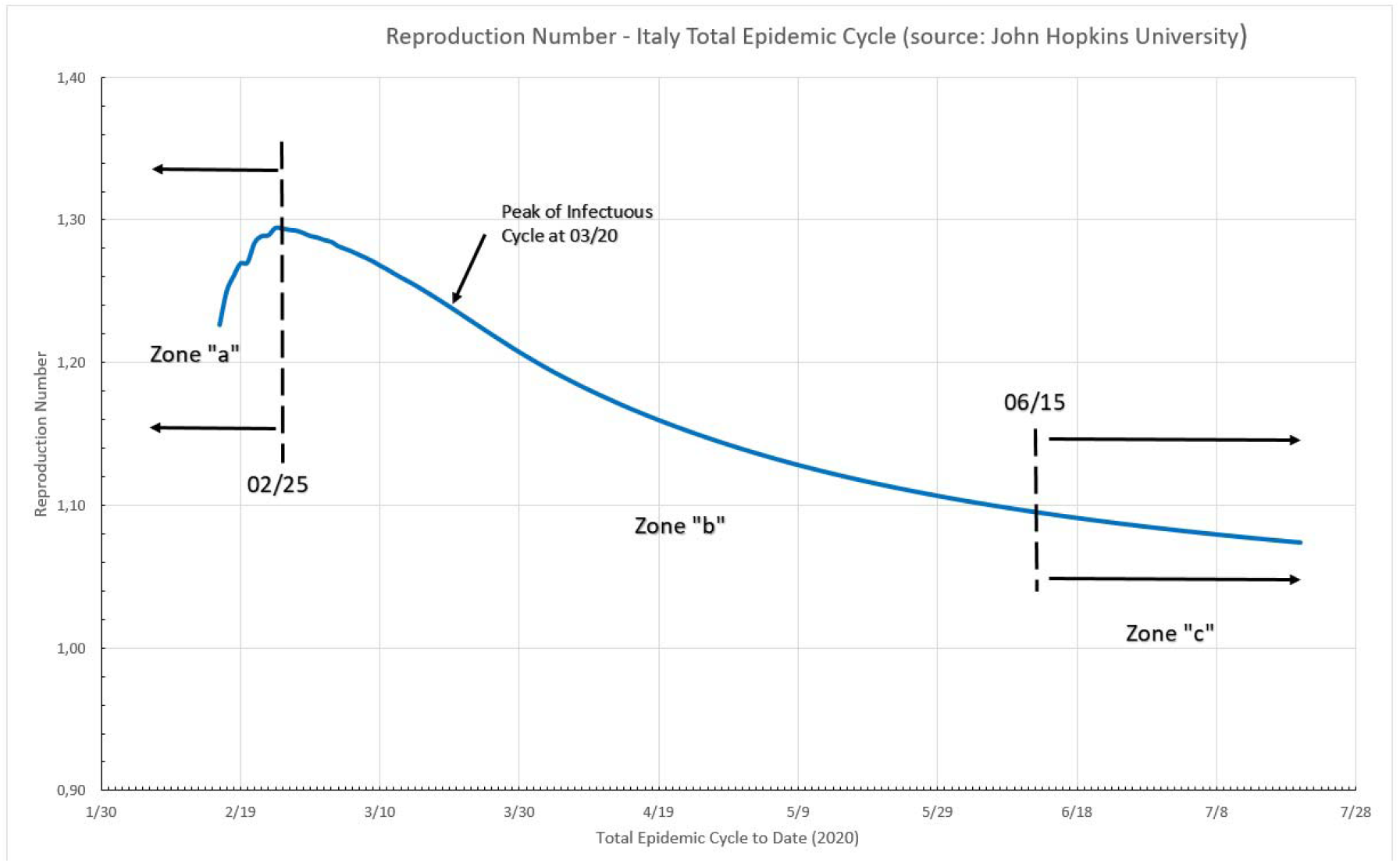
Total epidemic cycle in Italy, using the number of infected people daily.

**Figure 11.**
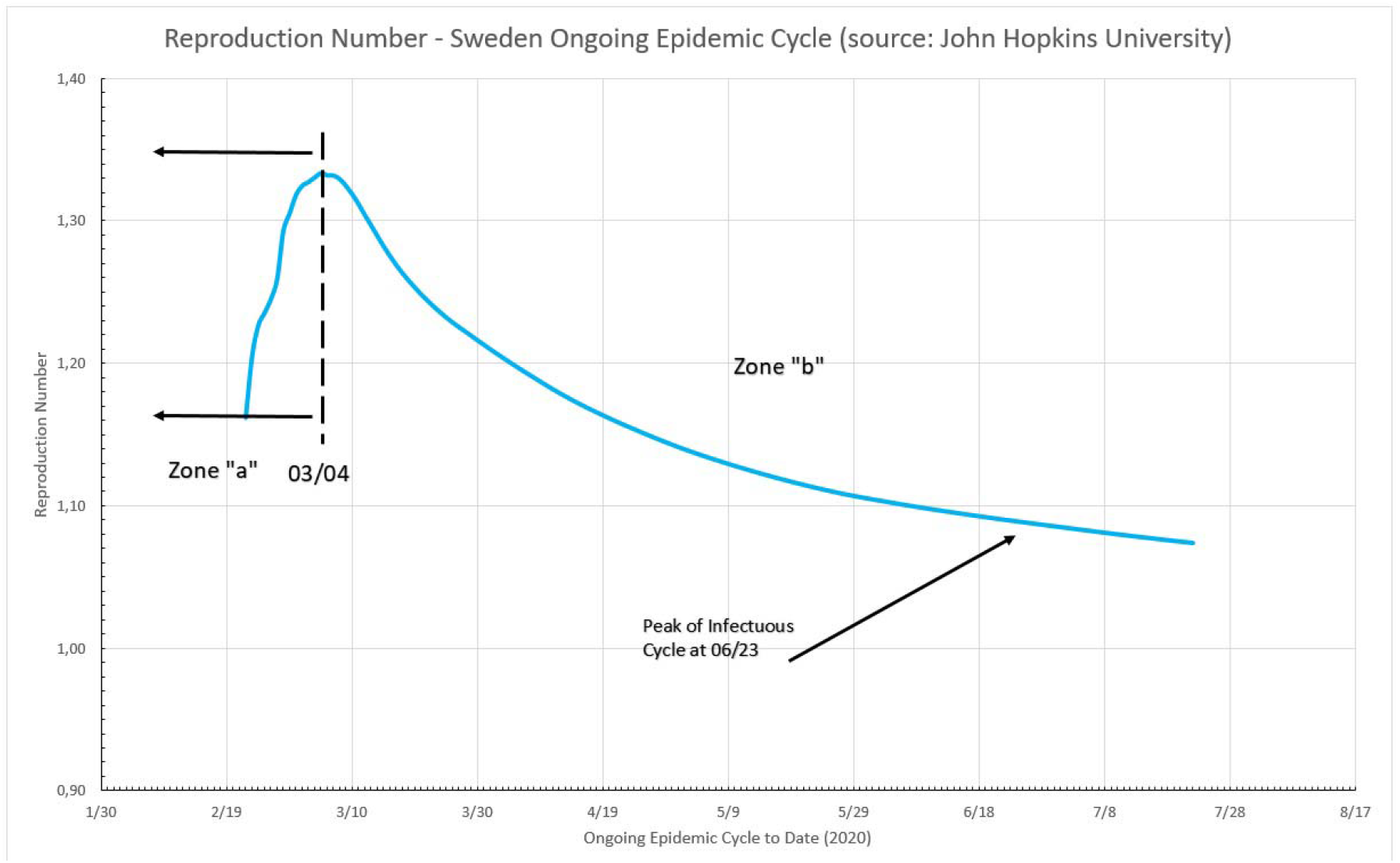
Ongoing epidemic cycle in Sweden, using the number of infected people daily.

**Figure 12.**
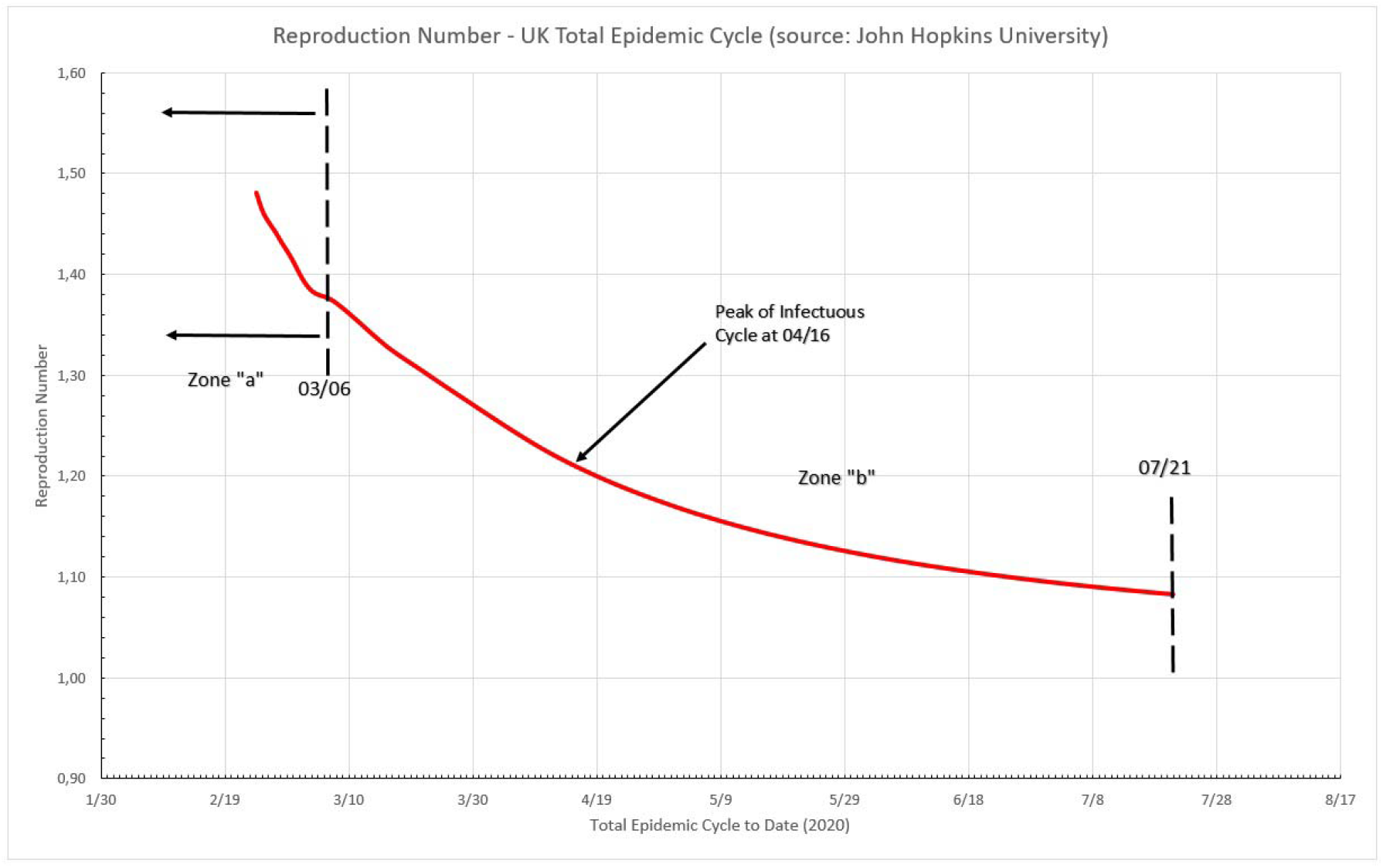
Total epidemic cycle in the UK, using the number of infected people daily.

### 4.5) South Korea

Data presented in Figure 13 shows two distinct zones, given that the registered cases starts exactly at the critical cycle. Zone “b” is in the beginning of the cycle and the Reproduction Number starts at 2.07 and drops very fast to 1.02. This is the critical disease cycle (from February 19^th^ up to April, 9^th^). After the limit date for the critical cycle, it is considered that Zone “c” is reached, which is the residual cycle.

**Figure 13.**
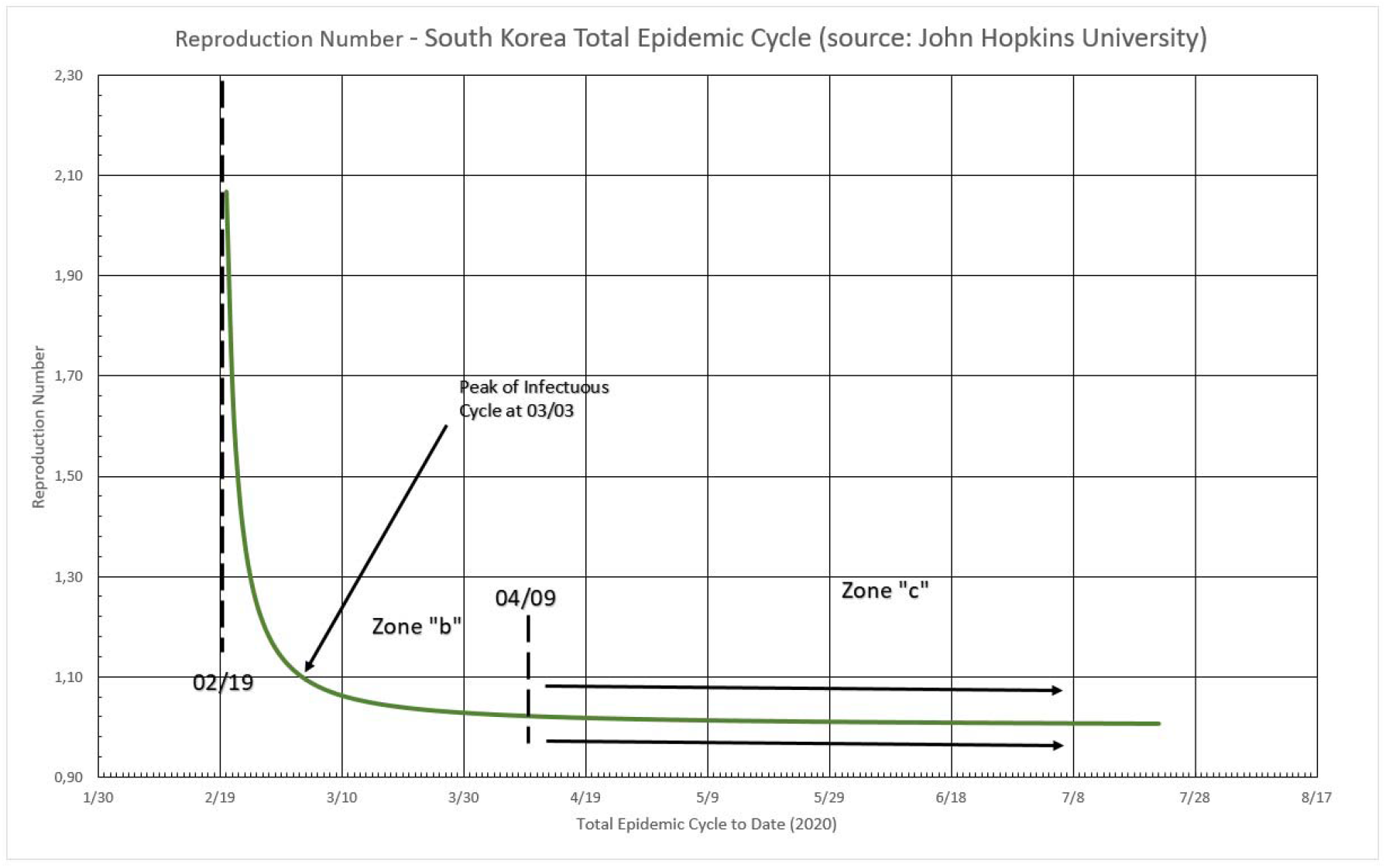
Total epidemic cycle in the South Korea, using the number of infected people daily.

### 4.6) New York State

In figure 14 two distinct zones are formed, because for the NY State, the critical cycle is still ongoing at the data collection day. Zone “a” is in the beginning of the cycle and the Reproduction Number oscillates between 3.94 and 3.30 from one day to the next, once again this probably is just the reflection of large initial variation in number, but this zone is limited to no more than 5% of the MAMI peak value. Zone “b” describes transmissivity during the critical disease cycle (from March 16^th^ onwards). This is the most lethal period of the epidemic cycle and it is considered over once a below 5% peak level is reached again. In absolute values the Reproduction Number for the critical period starts with a 3.30 value and drops continuously towards 1.08.

**Figure 14.**
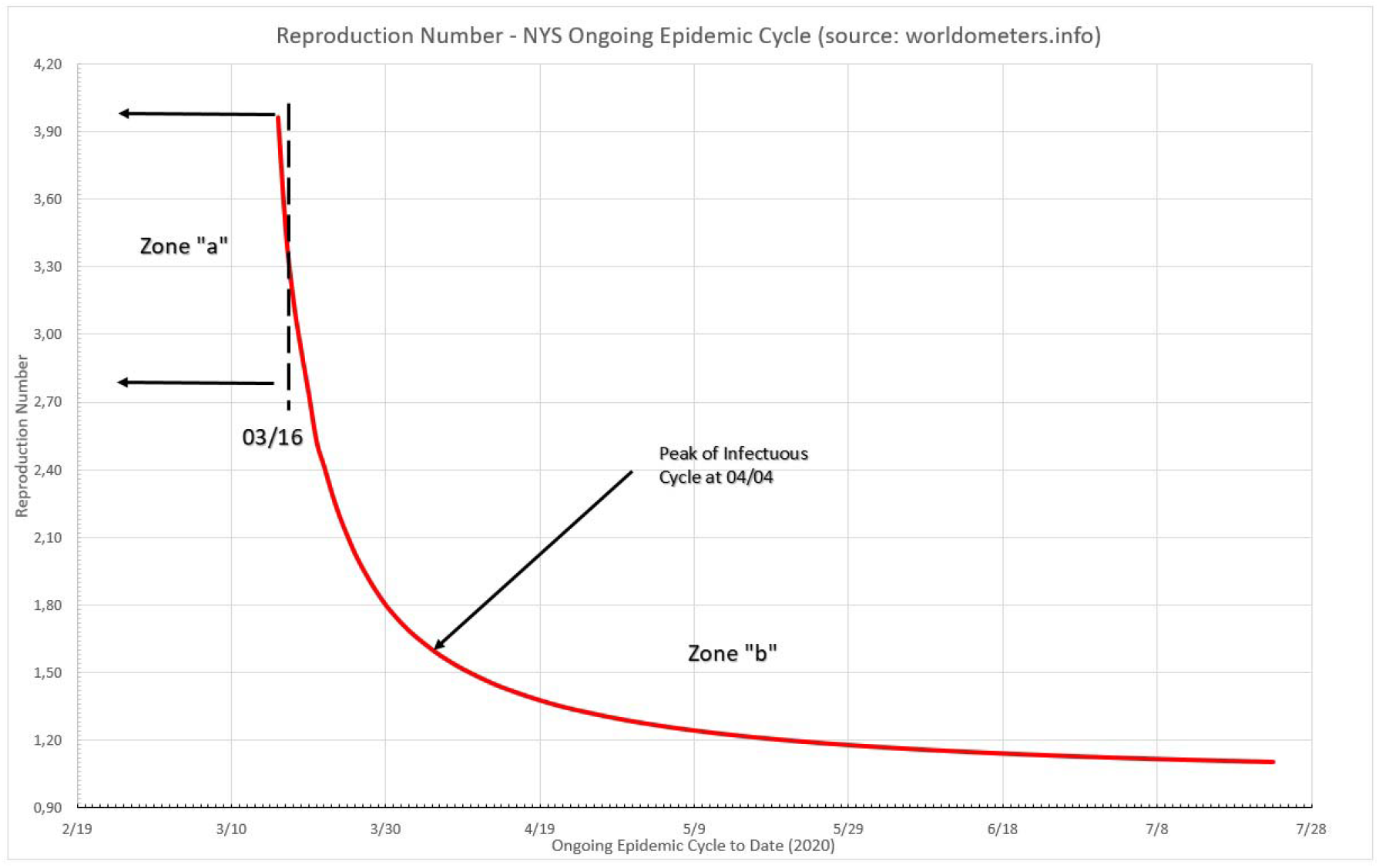
Total epidemic cycle in New York State, using Worldometers database as source for number of infected people daily.

## 5) Sub Notification Effect Over Reproduction Number

When it comes to analyzing the number of cases of infection in the COVID-19 epidemic, an issue that always arises is underreporting or sub-notification and its importance in predicting the behavior of the epidemic cycle. Thus, the second part of this work is dedicated to the study of sub-notification and its effects on prediction.

Sub-notification (SN) is here understood as the fact that the COVID-19 true number of infected are only estimated by public health authorities. Given that most of the people exposed to the virus does not display any sign of infection or the symptoms are very mild, therefore going unnoticed and unregistered by local bureaus of health statistics, the development of evaluation tools of the impact of these non notified cases is necessary.

If it is assumed that SN is a constant factor (eg. 10 times the registered number of cases) during the whole epidemic cycle, it does not change the absolute daily increase rate “b” or the (1 + b) factor. However, in fact it does affect the scale factor “a”, therefore changing the reproduction number (R_t_).

### 5.1) Sub Notification Impact Estimation Method

The impact of SN over R_t_ may be estimated by the following method. Initially, it is assumed that the actual registered numbers for daily contaminated people are no longer their actual values, but “real” ones multiplied by a factor, the SN factor. After that the scale factor “a” is calculated. The term (1 + b) remains constant, once the ratio (see term definition in Formula 3) remains constant. Then “a” and (1 + b) are applied to Formula 5, thus recalculating R_t_, now reflecting the effect of the imposed SN factor. This new R_t_ value would have been the correct one, in case all sub notified cases were suddenly registered. The percentage difference between this new, recalculated R_t_, and the actual one, provides an estimate for the impact of SN over the reproduction number for a given population. Therefore, multiplying the values for registered cases by a factor of 10 will not cause a tenfold increase in R_t_. The true impact must be, therefore, calculated as described. It is also observed that SN affects mostly at the very beginning of the critical cycle. After a certain amount of time, error drops to insignificant values (below 5%). Practical applications of this method are presented as follows.

### 5.2) Sub-Notification Estimates for Germany

An arbitrary threshold line representing a 5% error was drawn in Figure 15. This limit tells that after the 50^th^ day into the German critical cycle (the one between 5% of the peak value, before and after it), regardless the amount of sub notification, the error over the calculated Reproduction Number is no greater than 5%. At the other extreme, a 3x sub-notification basically does not induce errors larger than 5% over the Reproduction Number, in any time during the critical cycle. A maximum error of 16.84% is estimated for the worst case scenario here simulated, a 40x sub-notification, and the first day into the cycle. In overall, sub-notification appears to have no significant impact in Germany official infected numbers. Sub-notification also seems to have more impact in the very beginning of a given cycle, but turns itself irrelevant towards the end.

**Figure 15.**
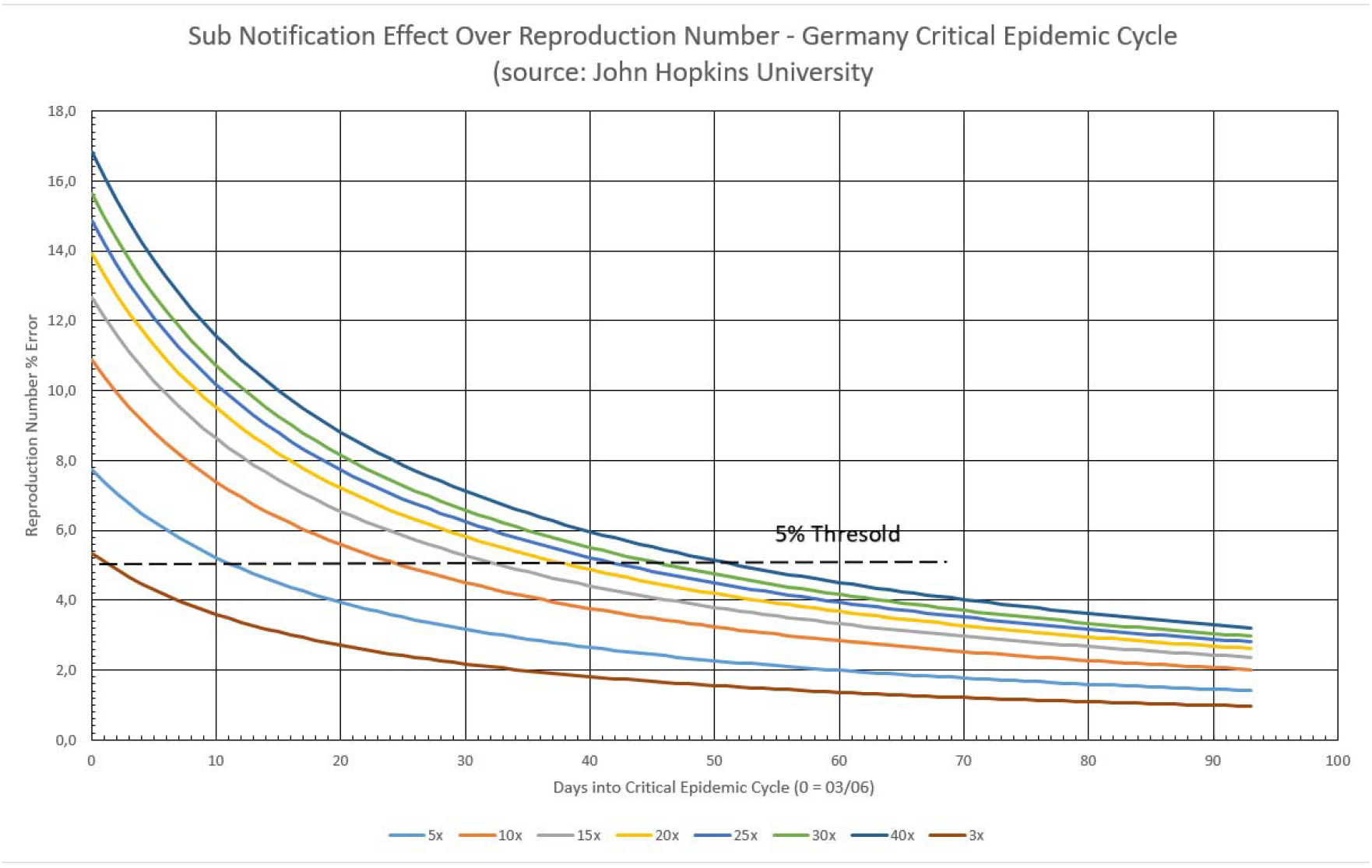
Sub notification effect over Transmissivity Rate in Germany, during the critical epidemic cycle.

### 5.3) Sub Notification Estimates for South Korea

The same threshold line representing 5% error was drawn in Figure 16. This limit tells that after the 48^th^ day into the South Korean critical cycle, regardless the amount of sub notification, the error over the calculated Reproduction Number is no greater than 5%. At the other extreme, a 3x sub notification basically does not induce errors larger than 5% over the Reproduction Number, in any time during the critical cycle. A maximum error of 14.25% is estimated for the worst case scenario here simulated, a 40x sub notification, and the first day into the cycle. In overall, sub notification appears to have no significant impact in South Korea, as in Germany, official infected numbers. Sub notification also have more impact in the very beginning of a given cycle, but turns itself irrelevant towards the end of it.

**Figure 16.**
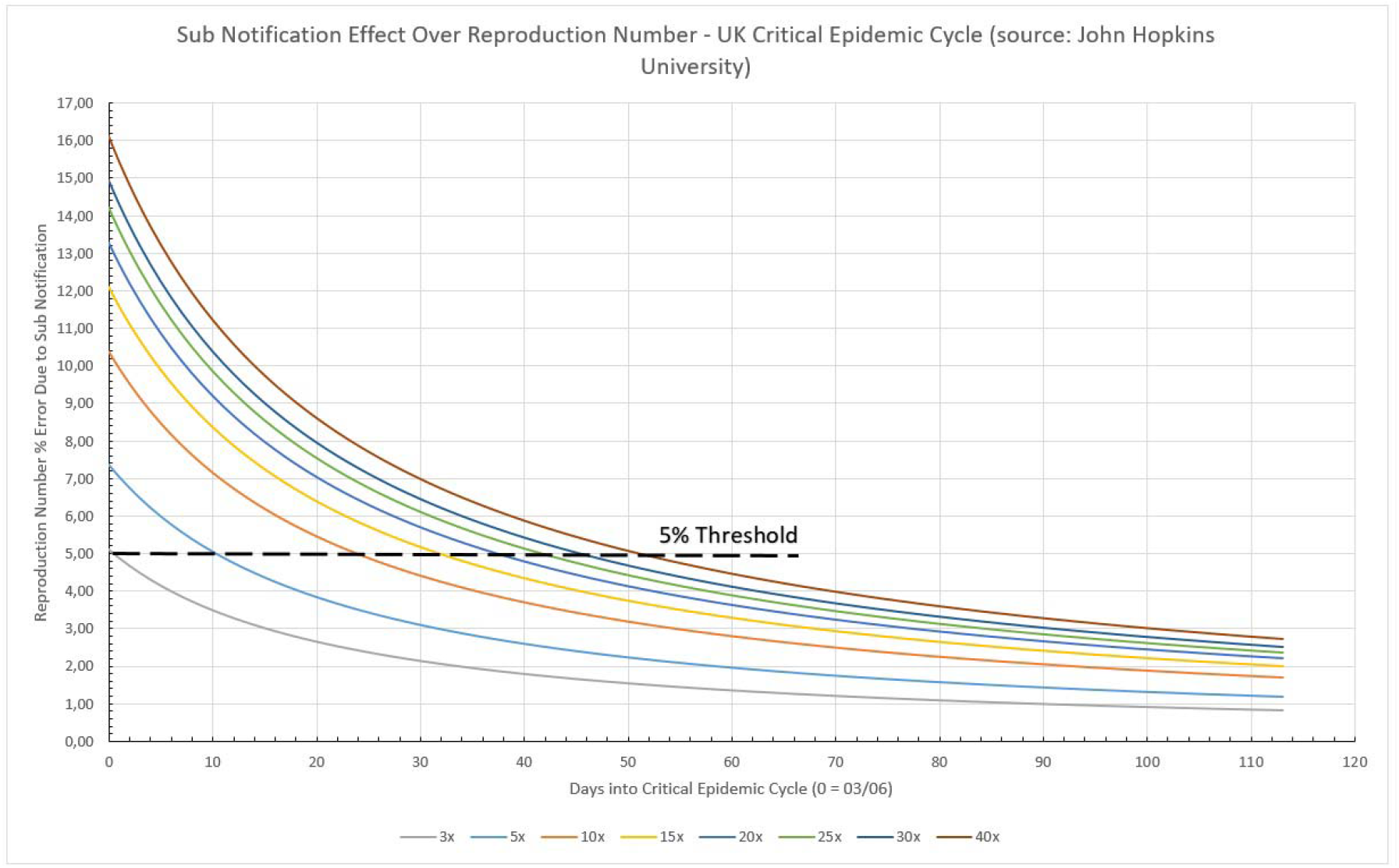
Sub notification effect over Transmissivity Rate in South Korea, during the critical epidemic cycle.

### 5.4) Sub Notification Estimates for Italy

Italy case is presented in Figure 17. The 5% limit tells that after the 44^th^ day into the Italian critical cycle, regardless the amount of sub notification, the error over the calculated Reproduction Number is no greater than 5%. At the other extreme, a 3x sub notification basically does not induce errors larger than 5% over the Reproduction Number, in any time during the critical cycle and 5x barely disturbs it. A maximum error of 12.34% is estimated for the worst case scenario here simulated, a 40x sub notification, and the first day into the cycle. In overall, sub notification appears to have no significant impact in Italy, as in the previous two cases, official infected numbers. Sub notification also have more impact in the very beginning of a given cycle, but turns itself irrelevant towards the end of it.

**Figure 17.**
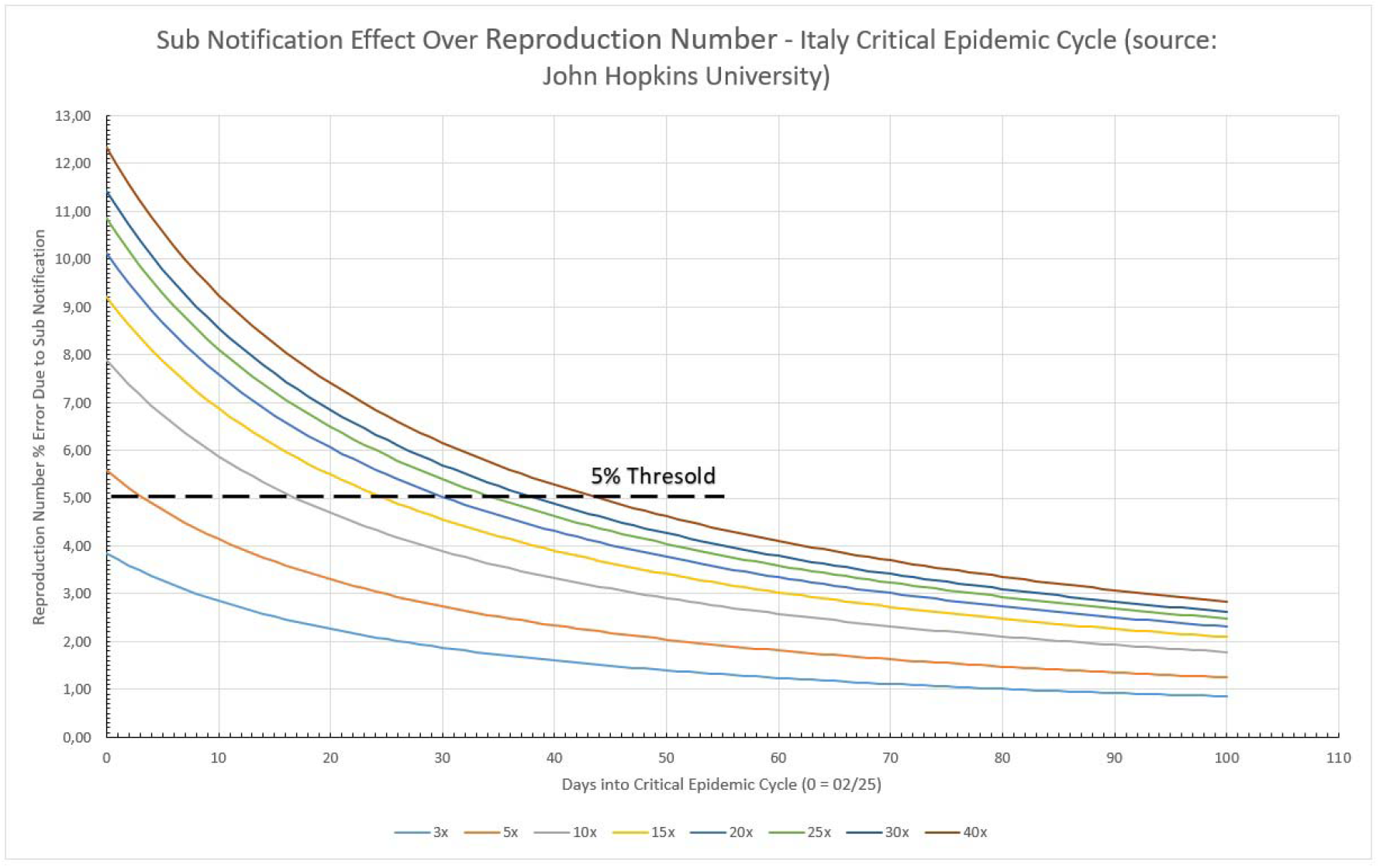
Sub notification effect over Transmissivity Rate in Italy, during the critical epidemic cycle.

### 5.5) Sub Notification Estimates for Sweden

Sweden SN effect is presented in Figure 18. The calculated limit tells that after the 54^th^ day into the Swedish critical cycle, regardless the amount of sub notification, the error over the calculated Reproduction Number is no greater than 5%. At the other extreme, a 3x sub notification basically does not induce errors larger than 5% over the Reproduction Number, after the fourth day during the critical cycle. A maximum error of 18.53% is estimated for the worst case scenario here simulated, a 40x sub notification, and the first day into the cycle. In overall, sub notification appears to have no significant impact in Sweden. Sub notification also have more impact in the very beginning of a given cycle, but turns itself irrelevant towards the end of it.

**Figure 18.**
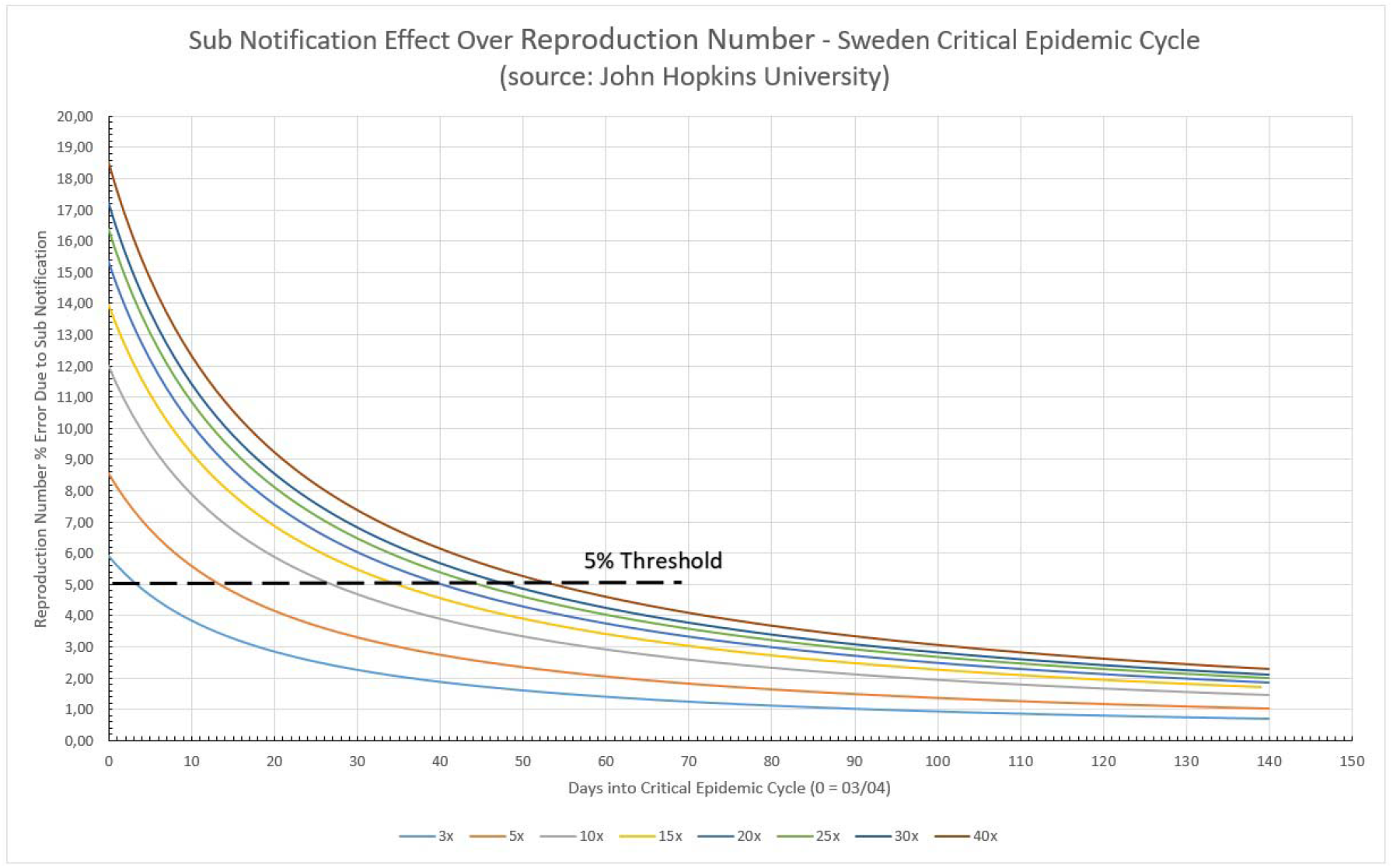
Sub notification effect over Transmissivity Rate in Sweden, during the critical epidemic cycle.

### 5.6) Sub Notification Estimates for New York State

NY State case is presented in Figure 19. Its limit tells that after the 63^rd^ day into the New York critical cycle, regardless the amount of sub notification, the error over the calculated Reproduction Number is no greater than 5%. At the other extreme, even at a 3x sub notification, the error induced is much larger than 5% over the Reproduction Number, and takes 13 days to reach values below 5%. A maximum error of 33.63% is estimated for the worst case scenario here simulated, a 40x sub notification, and the first day into the cycle. In overall, sub notification appears to have some impact in NY State. Sub-notification also have more impact in the very beginning of a given cycle, but turns itself irrelevant towards the end of it.

**Figure 19.**
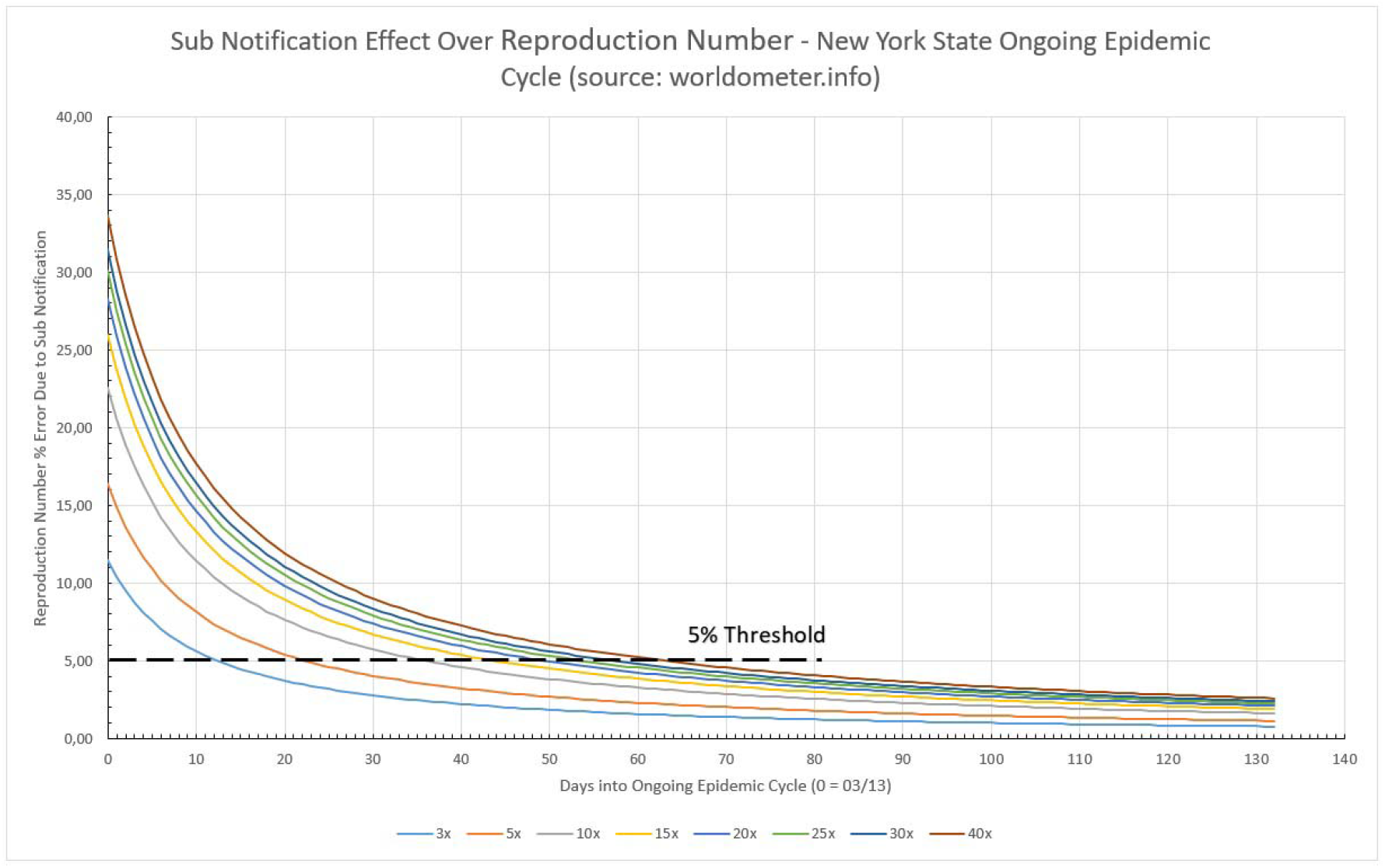
Sub notification effect over Transmissivity Rate in NY State, during the critical epidemic cycle.

### 5.7) Sub Notification Estimates for the United Kingdom

UK case is presented in Figure 20. The 5% limit tells that after the 51^st^ day into the British critical cycle, regardless the amount of sub notification, the error over the calculated Reproduction Number is no greater than 5%. At the other extreme, a 3x sub notification basically does not induce errors larger than 5% over the Reproduction Number, in any time during the critical cycle and 5x barely disturbs it and only during the first 10 days. A maximum error of 16.11% is estimated for the worst case scenario here simulated, a 40x sub notification, and the first day into the cycle. In overall, sub notification appears to have no significant impact in Italy, as in the previous 2 cases, official infected numbers. Sub notification also have more impact in the very beginning of a given cycle, but turns itself irrelevant towards the end of it.

**Figure 20.**
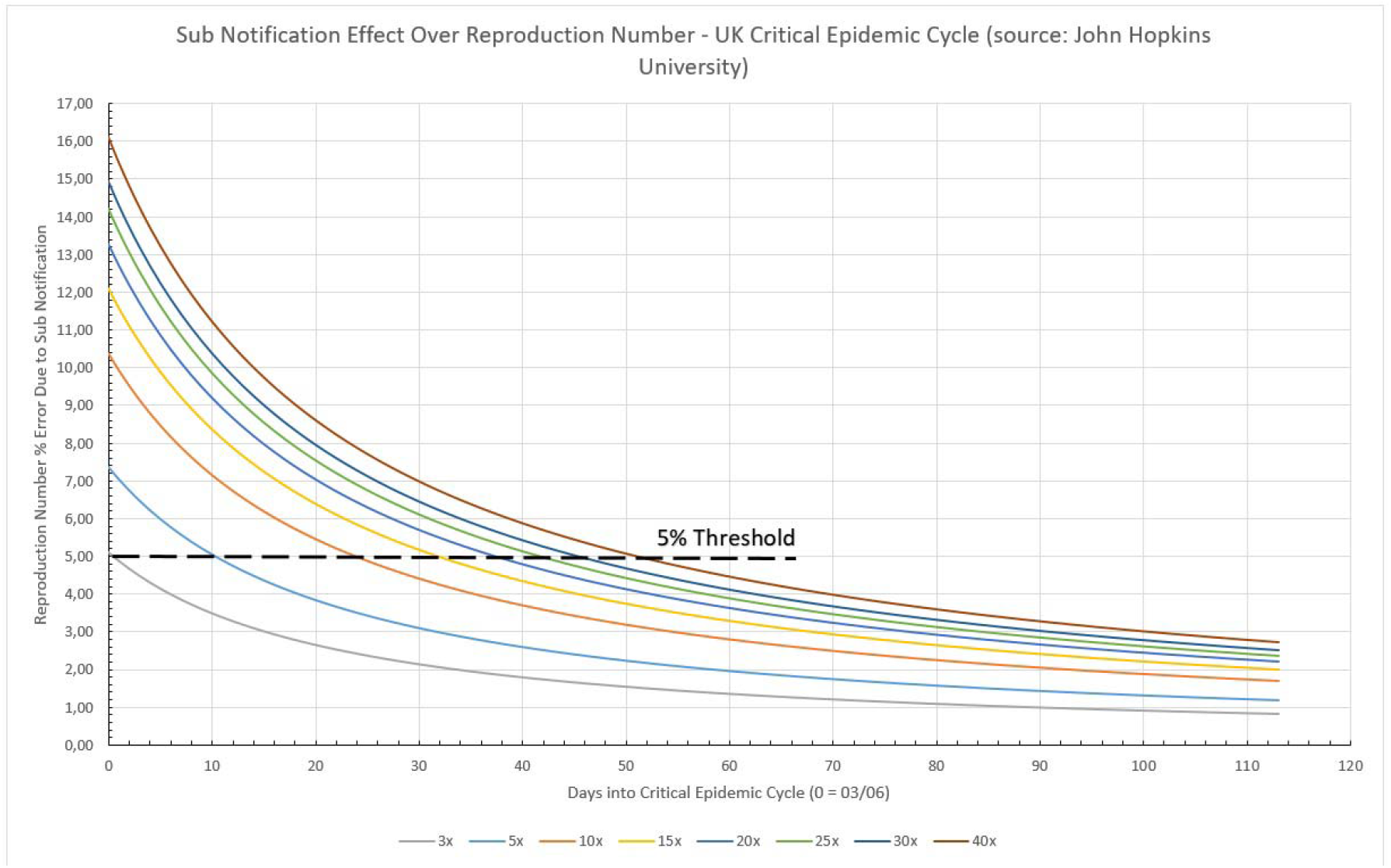
Sub notification effect over Transmissivity Rate in the UK, during the critical epidemic cycle.

## 6) Most Lethal Cycle of Epidemics Control Performance

This item expresses the epidemic behavior during the Most Lethal Cycle of Epidemics (MLCE) [1], which is defined as the period of time when the number of registered cases grows fast towards a peak and then declines towards an almost constant daily rate. The frequency shape resembles a triangular form, where the ascending side is smaller than the descending side [1].

In order to allow a comparison between different MLCEs all listed places had their date normalized. The interval was selected between the established 5% limits, and the non-dimensional time was determined dividing each day by the total MLCE duration, as listed in Table 1. For the Reproduction Number, all the values were divided by the largest value found in the MLCE interval. All these transformations allow to confront how efficient was the disease spreading control used. Sweden and New York State were considered as still having an open MLCE, therefore the end of the cycle considered was the day of the data collection (July, 22^nd^, 2020).

Figure 21 shows that Italy was, relatively, the most unsuccessful place in reducing Reproduction Numbers, although not for a large margin. Germany and the UK have the same performance and kept R_t_ falling slowly but steadily. South Korea and New York State achieved a large drop in the early stages of the critical cycle, but after that R_t_ were basically constant.

**Figure 21.**
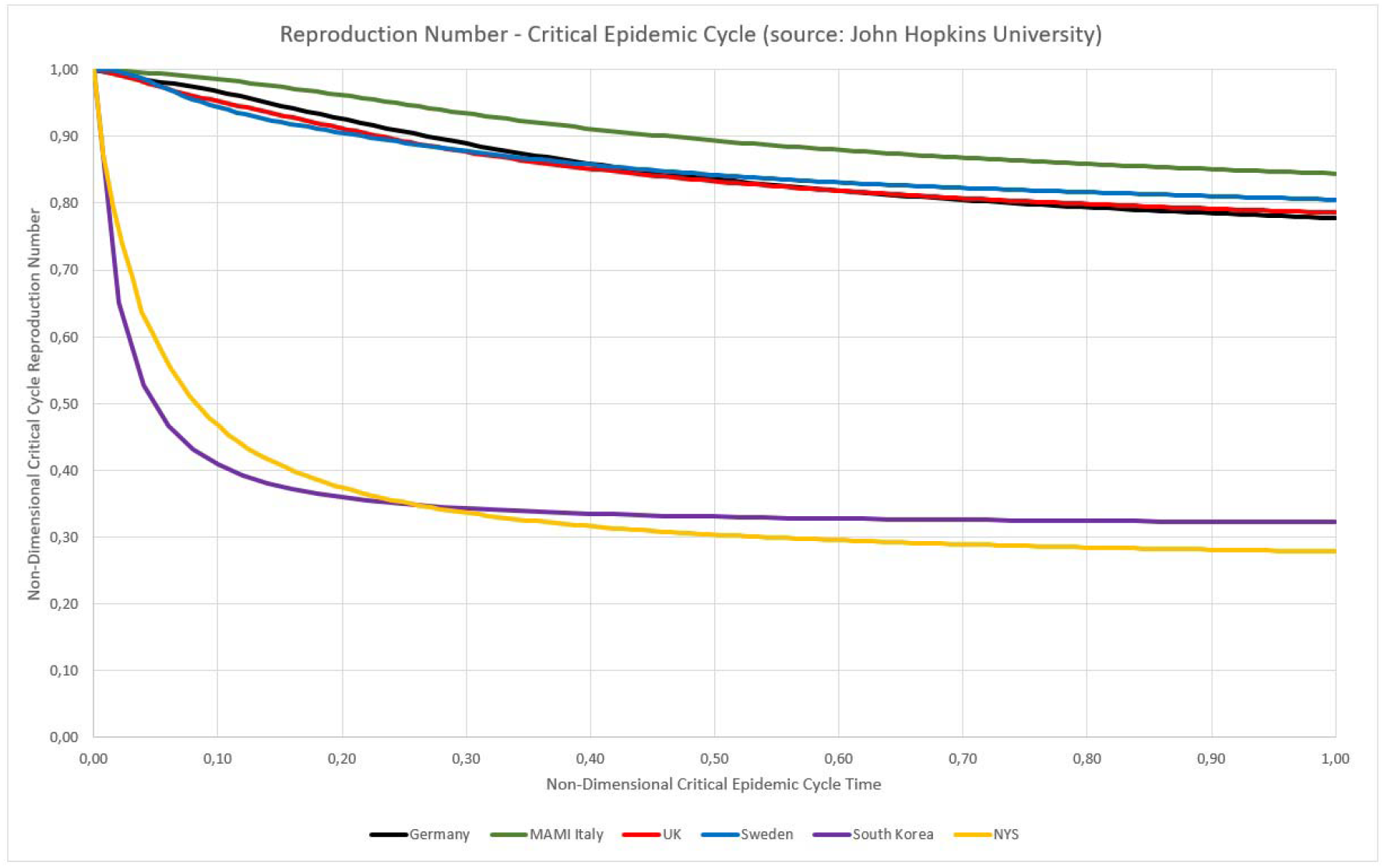
Non-dimensional critical epidemic cycle for Germany, Sweden, UK and Italy.

## 7) Conclusions

In an attempt to quickly predict the evolution of local episodes of COVID-19 in countries and cities, several prediction models for epidemic cycles were presented, most of them, unfortunately, were not successful, as the pandemic progressed and consequent access to updated data, as well as the literature, showed.

As a natural evolution of this task of prediction, new models emerged, adding complexity in the search for success in forecasts. Although some models have been successful, we understand that, in order to become usable by a significant number of health authorities, in addition to the necessary efficacy, they also need to be efficient, which in our view, is related to simplicity of use. Models that are too complex, even when supported by software, still need to be fed by several parameters, which makes these tools often inaccessible to decision makers in locations with less technical-scientific resources.

In this sense, this article complements the previous one [1], which dealt with prediction in the number of deaths, in a two fold way: by bringing an effective and efficient model of prediction in the number of infection cases, and by also seeking to show the impact of sub-notification on calculations, shedding light on this discussion, which is still frequent, especially in countries where there is no mass testing.

We show that, with relatively simple mathematical tools, and even in the face of data with quality problems, it is possible to obtain reliable values for time-dependent, incubation period-independent Reproduction Numbers (R_t_), as well as we demonstrate that the impact of sub-notification, when provided with the proper statistical treatment, is relatively low, after the initial phase of the epidemic cycle has passed. In this way, we believe to have contributed to the studies of the epidemic cycles of COVID-19.

## Data Availability

All data is available in the public sites of JHU and Worldometers on COVID-19. The dates in which their were retrieved are informed in the paper.

https://coronavirus.jhu.edu/map.html

https://www.worldometers.info/coronavirus/

